# Polyphenol metabolites in human milk: Potential role in support of healthy infant development, a narrative review

**DOI:** 10.1101/2025.02.04.25321667

**Authors:** Chelsey Fiecke, Nicole Knox, Aline Andres, Mario G. Ferruzzi, Colin Kay

**Affiliations:** Arkansas Children’s Nutrition Center and Department of Pediatrics, University of Arkansas for Medical Sciences, Little Rock, AR, 72202, United States

**Author notes:** **Correspondence: Colin Kay,** Arkansas Children’s Nutrition Center and Department of Pediatrics, University of Arkansas for Medical Sciences, Little Rock, AR, United States., **Mario Ferruzzi,** Arkansas Children’s Nutrition Center and Department of Pediatrics, University of Arkansas for Medical Sciences, Little Rock, AR, United States. Department of Food Science, College of Agriculture and Life Sciences, Virginia Polytechnic Institute and State University, Blacksburg, VA, 24061, United States.

**Keywords:** polyphenol, human milk, cognitive development, maternal diet, lactation

## Abstract

Polyphenols are a broad class of phytochemicals derived from healthy diets rich in fruits, vegetables, and whole grains. Due to emerging evidence linking healthy dietary patterns to maternal and child health benefits, there is increasing interest in dietary bioactives such as polyphenols in human milk. This narrative review critically evaluates evidence supporting the transfer of polyphenols to breastfed infants in human milk, potential role of maternal diet and other non-dietary characteristics (e.g., lactation duration, genetics) in modifying polyphenol composition and concentrations in human milk, and the relationship between human milk polyphenols and infant development. Over the last decade, 20 studies have reported polyphenols or phenolic metabolite concentrations in human milk. Although polyphenols can be transferred to infant circulation via human milk, evidence supporting a role for human milk polyphenols in infant development is still limited. Maternal dietary intake influences polyphenol composition and concentrations of human milk, with other reports suggesting that maternal metabotype, lactation stage, demographics, and genetics may play a role. Future studies need to include a broader assessment of polyphenols in human milk, and systematic integration of additional maternal factors that may influence polyphenol metabolism or human milk composition.

## 1. Introduction

Critical periods of neurodevelopment occur during the first 1,000 days of life (conception to 2 years old), with nutritional exposures impacting both short- and long-term cognitive function [1,2]. Increasing child exposure to bioactive components from prenatal maternal diet and human milk (e.g., fruit and vegetable consumption) have been associated with enhancement of cognitive outcomes in offspring [3–6]. Benefits of breastfeeding stem from the unique composition of human milk, which includes a growing list of bioactives associated with healthy infant growth and development [7].

The most studied human milk bioactives include immunoglobulins, hormones, proteins, human milk oligosaccharides (HMOs), white blood cells, peptides, cytokines, chemokines, and miRNA [8]. More recently, the Breastmilk Ecology: Genesis of Infant Nutrition (BEGIN) Project, led by the National Institute of Child Health and Human Development (NICHD), has expanded this list to also include dietary bioactives derived from maternal intake of plant-based foods [9]. These bioactives include the omega-3 fatty acids such as docosahexaenoic acid (DHA) and α-linolenic acid, the carotenoid lutein, and polyphenols, which are provided by healthy dietary patterns rich in fruits, vegetables, whole grains, and fish [10–12], and can be transferred to infants through human milk [13–16]. In the case of DHA, specific functional roles, particularly in supporting visual [17] and cognitive development [18,19], have led to expert scientific organizations such as the Food and Agriculture Organization [20] to recommend a nutrient intake level of 200 mg/d during pregnancy and lactation. Similar efforts to frame recommendations for lutein intake during pregnancy and lactation are underway [16,21] due in large part to evidence supporting a role for lutein in ocular [22–24] and brain maturation [19,25,26].

Unlike lutein and DHA, which are singular compounds derived from specific foods and have a relatively long history of study in human milk, polyphenols have only recently been identified as components of human milk [27–30]. Polyphenols are a broad class of diet-derived bioactives, with exposure primarily from consumption of plant-based foods and beverages containing fruits, vegetables, cocoa, tea, and coffee. Daily intakes for polyphenols in the U.S. adult population can reach up to 1,200-1,900 mg/d. The potential health benefits of a polyphenol-rich diet have been well-documented in adults. Specifically, intake of polyphenols and polyphenol-rich foods has been linked to improvements in cardiovascular [31], cardiometabolic [32], and cognitive outcomes [33]. For adult populations, the associations between flavonoids, a subclass of polyphenols, and cardiovascular health benefits has resulted in recent recommendations from the Academy of Nutrition and Dietetics for levels of dietary intake in adults of 400-600 mg/d [34]. Clinical studies evaluating developmental impacts of polyphenols or polyphenol-rich dietary components due to exposures occurring during pregnancy, infancy, or childhood are limited to observational studies [3,4,35–38] and a few dietary interventions [39–41]. These studies demonstrate that fruit and vegetable consumption, a major source of polyphenol intake, in expectant women [3,4,36,38,39], infants [35,37], and children [40,41] positively impacts cognitive development and function that may last through adolescence.

With emerging evidence linking specific foods and diet patterns to maternal and child health benefits, interest in understanding infant exposure to bioactive components, such as polyphenols, from human milk has increased. Over the last decade, our group and others have reported the presence of physiologically relevant concentrations of polyphenols and their metabolites in human milk. As a part of the overall bioactive composition of human milk, it remains to be seen the extent to which these compounds may contribute to optimization of healthy infant growth and development. With this in mind, the goal of this narrative review is to critically evaluate existing evidence that supports: 1) the transfer of polyphenols and related metabolites to infants during breastfeeding, 2) the potential role that maternal diet and non-dietary factors (e.g., genetics, metabotype, demographics, and lactation duration) play in modulating human milk polyphenol composition, and 3) the relationship between polyphenol intake in breastfed infants and infant development. We also present a framework for future research priorities aimed at understanding if polyphenol profiles in human milk are linked to healthy infant development.

## 2. Materials and Methods

### 2.1. Search strategy

Pertinent studies were identified by searching PubMed and Google Scholar electronic databases using the following keywords: ‘flavonoid’, ‘flavan-3-ol’, ‘anthocyanin’, ‘catechin’, ‘epicatechin’, ‘epigallocatechin gallate’, ‘phenolic’, ‘phenolic compound’, ‘polyphenol’, or ‘isoflavone’, and ‘breast milk’ or ‘human milk’. Additional studies were identified from manual searches of reference lists in relevant original and review articles. A total of 83 studies were identified, with 63 studies being excluded. Included studies (n = 20) were peer-reviewed publications and presented original research [10,13,14,27–30,42–54]. Studies were excluded if they lacked quantified concentrations of individual phenolic compounds (41%), quantified only environmental contaminants or synthetic antioxidants (19%), reported duplicate data (5%), were derived from pre-clinical models (21%), not original research published in English (12%), or published only in abstract form (3%; e.g., conference proceedings) (**Table 1**; inclusion and exclusion criteria).

**Table 1.** Inclusion and exclusion criteria for literature search.

| Inclusion criteria | Exclusion criteria |
| --- | --- |
| Peer-reviewed publications in English | Review articles or expert panel reports |
| Original research including pharmacokinetics in human participants, dietary or supplement interventions in human participants, and observational cohort studies | Published only in abstract form |
|  | Lack of reported quantified individual phenolic concentrations in human milk (e.g., reliance on non-specific or indirect methodology such as total polyphenol content, total antioxidant capacity) |
|  | Pre-clinical studies (e.g., *in vitro* or animal models) |
|  | Analysis of phenolic compounds derived from environmental contaminants (e.g., PCBs, BPA) or synthetic antioxidants (e.g., BHT) |
|  | Duplicate data reported |
| PCBs, polychlorinated biphenyls; BPA, bisphenol A; BHT, butylated hydroxytoluene |  |

Throughout this manuscript the term polyphenol is used to refer to (poly)phenols. It is important to clarify that the term “(poly)phenol refers to a several phytochemical subclasses, including flavonoids, lignans, stilbenes, and phenolic acids. For clarity throughout, we used polyphenol to refer to this collective of phytochemical subclasses.

## 3. Results

### 3.1. Transfer of polyphenols and related metabolites to infants during breastfeeding

#### 3.1.1. Overview of polyphenols in food and metabolites present in circulation and human milk

Polyphenols are secondary plant metabolites [55] with a wide range of chemical structures [55] and physiological activities [19,56–60]. Over 8,000 polyphenolic compounds have been documented [61] in plant-based foods such as vegetables [62], fruits [63], spices [64], grains [65], legumes [66], nuts [67], coffee [68], and tea [69]. Major classes and subclasses of polyphenolic compounds and their common food sources are shown in **Figure 1** [70]. Quantification of dietary polyphenol intake in lactating women is scarce but has been reported recently in Argentinian women [71], which reported a total polyphenol intake is ∼1,000 mg/d, which is slightly lower than the average intake for non-lactating women in the U.S. (∼1,500 mg/d) [72]. Major polyphenols consumed by lactating women and common food sources are shown in **Table 2** [71,73].

**Figure 1:**
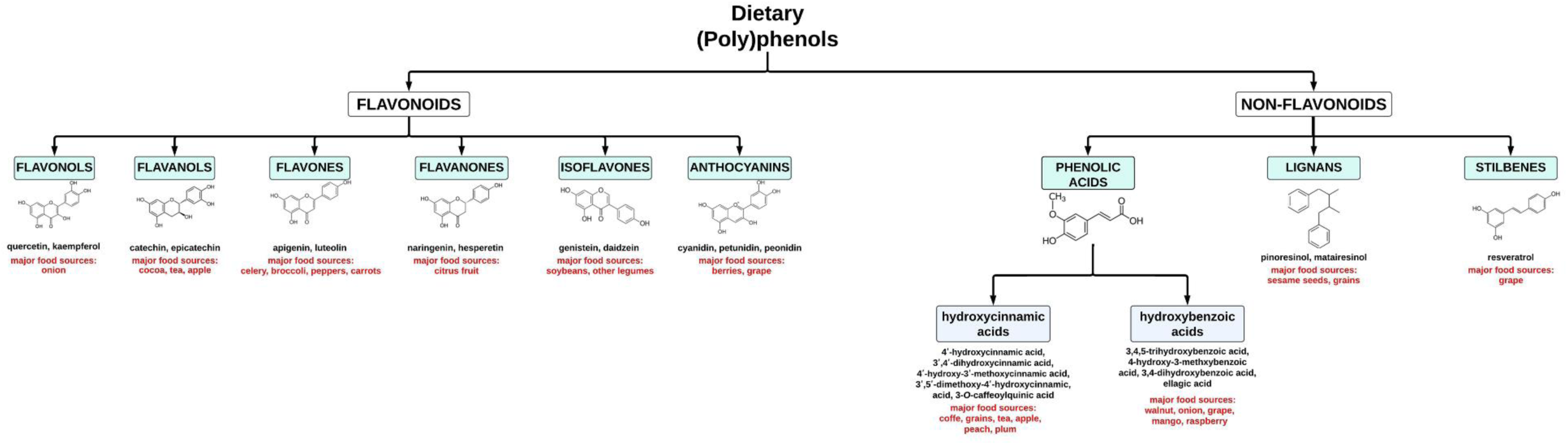
Major classes and subclasses of polyphenols, common food sources, and corresponding chemical structures. Adapted from Meccariello. [70].

**Table 2.**
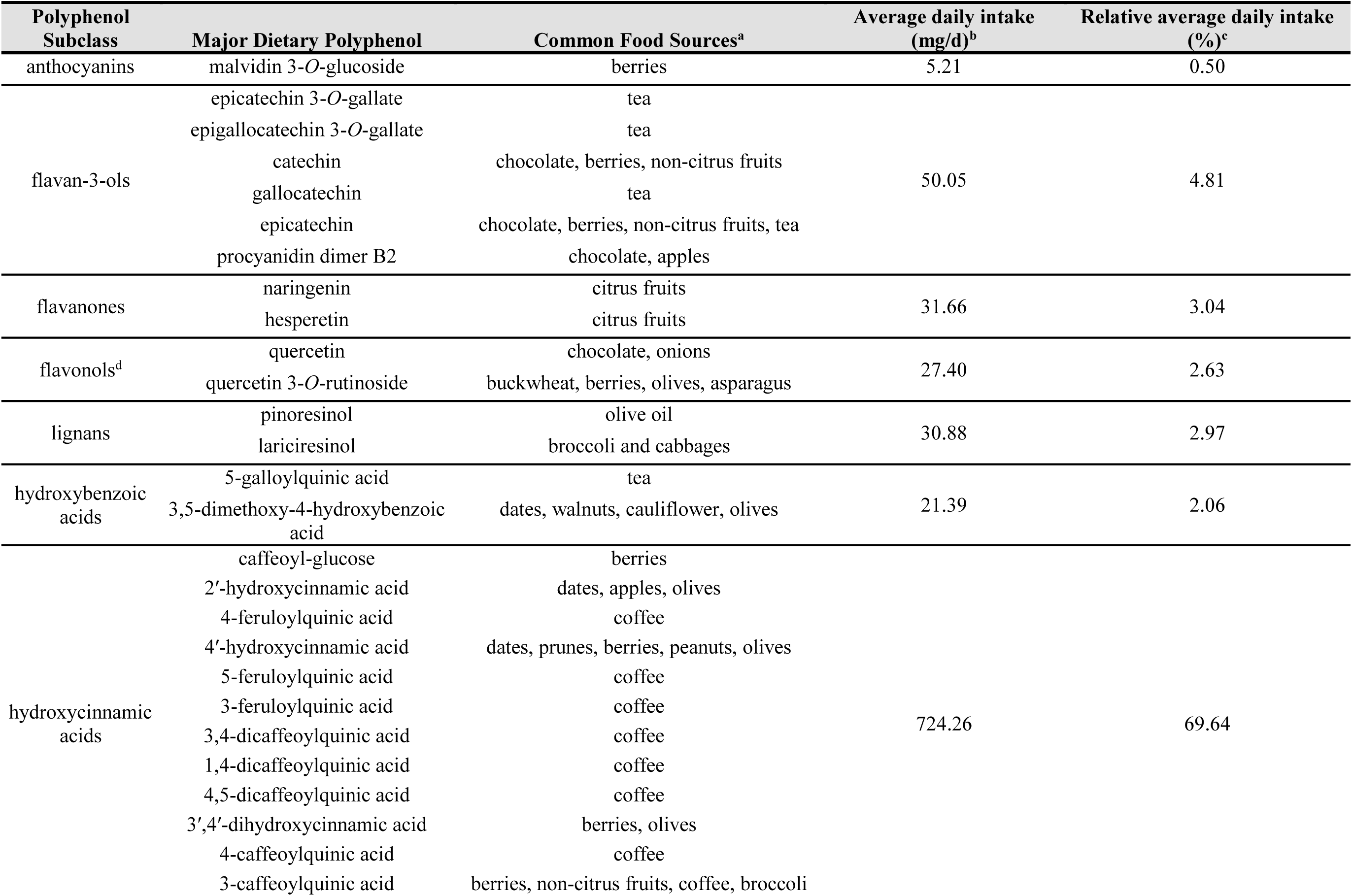
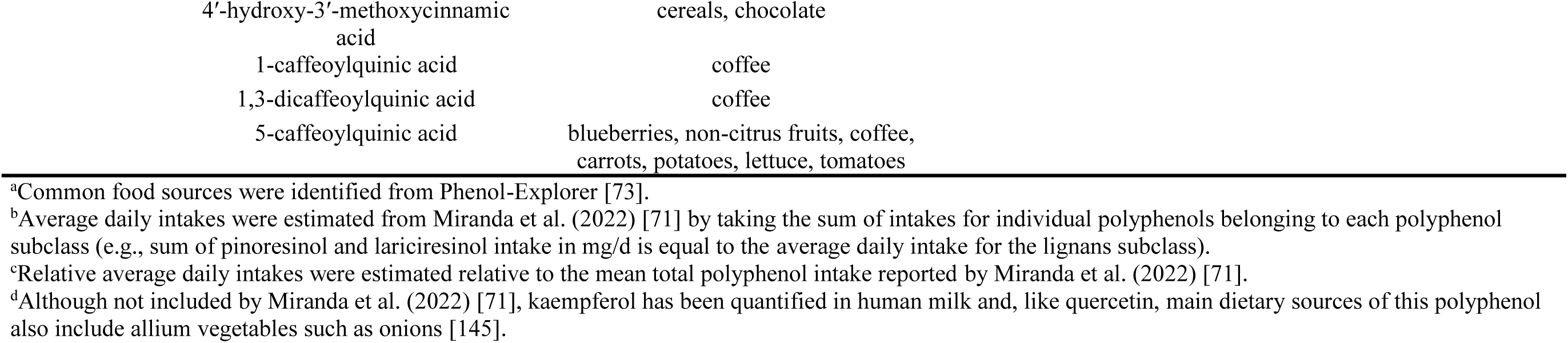
Major dietary polyphenols consumed by lactating women (>5 mg/d) and food sources.

Bioavailability of flavonoids and phenolic acids is considered poor in the upper gastrointestinal tract [74–77] due to size, polarity, and attached sugar moieties (e.g., flavonoid glycosides) and organic acids (e.g., chlorogenic acids). Additionally, phenolic acids can be found in nature bound to cell wall polysaccharides [74], which also limits their bioavailability in the upper gastrointestinal tract. Flavonoid glycosides can be hydrolyzed to aglycones by lactase phlorizin hydrolase (LPH) at the brush border of small intestine epithelial cells [78] or by microbes along the length of the gastrointestinal tract [79]. In adults, LPH expression occurs primarily in the jejunum, while both localization of LPH and degree of expression along the small intestine (duodenum, jejunum, and ileum) are enhanced during early postnatal life prior to introducing complementary foods [80], which may impact polyphenol metabolism and absorption in infants. Flavonoids and phenolic acids not absorbed in the small intestine pass into the lower gastrointestinal tract where microbial metabolism can lead to removal of conjugated moieties [81–83] and ring fission, allowing absorption of the aglycone form. Flavonoids and phenolic acids not absorbed in the small intestine pass into the lower gastrointestinal tract where microbial metabolism can lead to removal of the phase II conjugated moieties [81–83], allowing absorption of the aglycone form and/or further ring fission of polyphenols to small molecule phenolic metabolites [84,85], leading to production of various microbial metabolites such as hydroxyphenylacetic acids, hydroxyphenylpropanoic acids, urolithins, valerolactones, and short-chain fatty acids. After absorption of these microbial metabolites by the host, sulfation, glucuronidation, and/or methylation may occur by phase II metabolism in the intestine or liver [86]. As a result, various metabolites derived from dietary polyphenol intake may be present in circulation, including the intact food-derived compounds as well as host (phase II) and microbial metabolites, with those of microbial origin representing the greatest proportion [87].

Major types of polyphenols and host and microbial metabolites present in circulation are summarized in **Figure 2**. Polyphenols reported in human milk include dietary polyphenols [30] and phenolic metabolites [28,30] that are absorbed from dietary intake and subsequently transferred from circulation to human milk [13,14,42]. For infants under the age of six months, human milk provides the major dietary source of polyphenols due to ∼10-fold lower concentrations (10 mg/L) in infant formula [88] compared to human milk (95 mg/L) [89].

**Figure 2:**
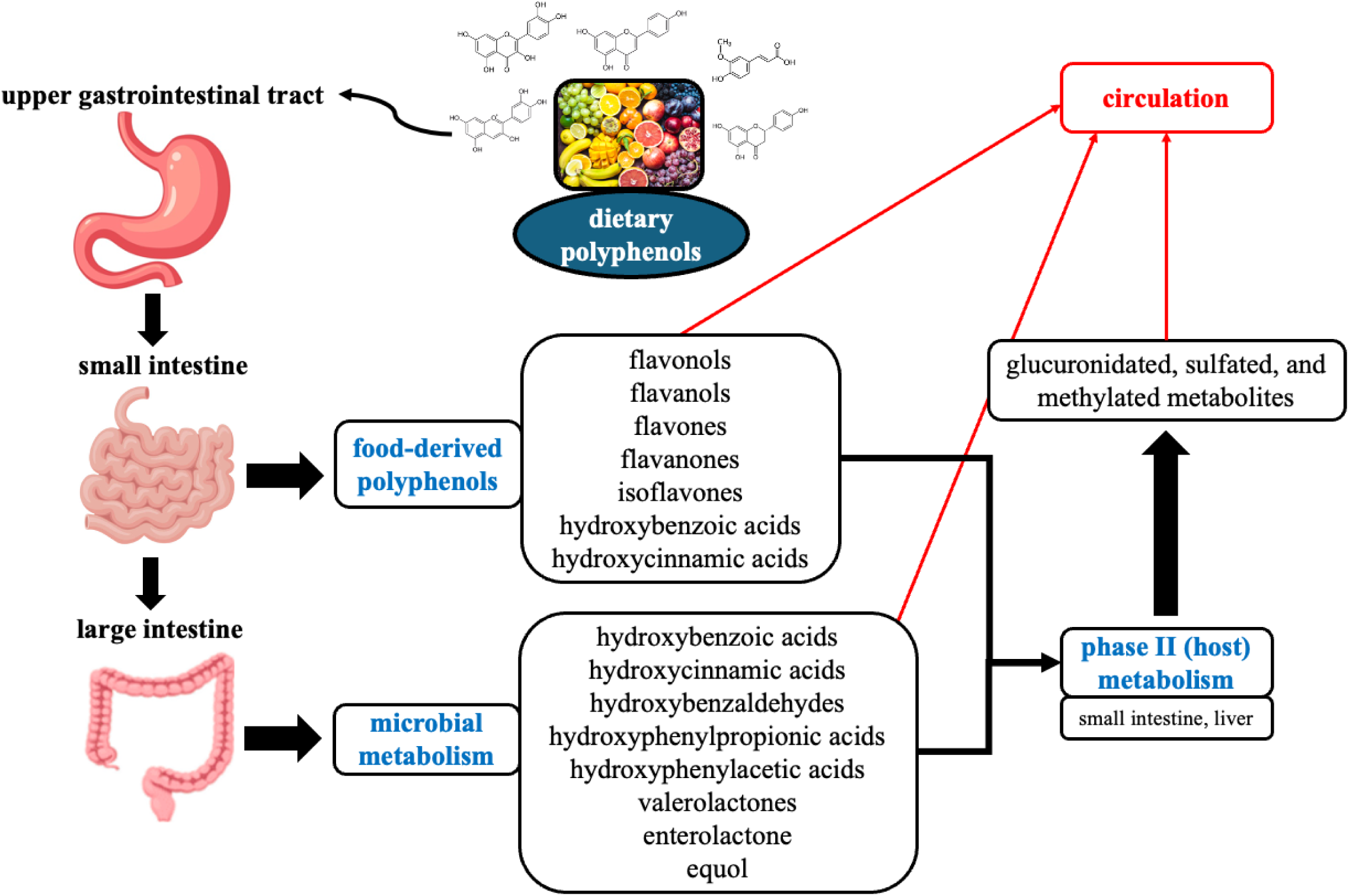
Major polyphenols present in circulation, including food-derived parent compounds and metabolites derived from host (phase II) and microbial metabolism.

#### 3.1.2. Infant exposures to polyphenolic compounds and metabolites in human milk

To date, there are 20 reports of polyphenols and phenolic metabolites in human milk, including single-dose pharmacokinetic studies [14,42,45–47], short duration dietary interventions (≤ 4 weeks) (**Fig. 3**) [13,29,43,44,48], and observational cohorts of free-living lactating women [10,27,28,30,49–54]; study characteristics are summarized in **Table 3** and **Table 4**.

**Figure 3:**
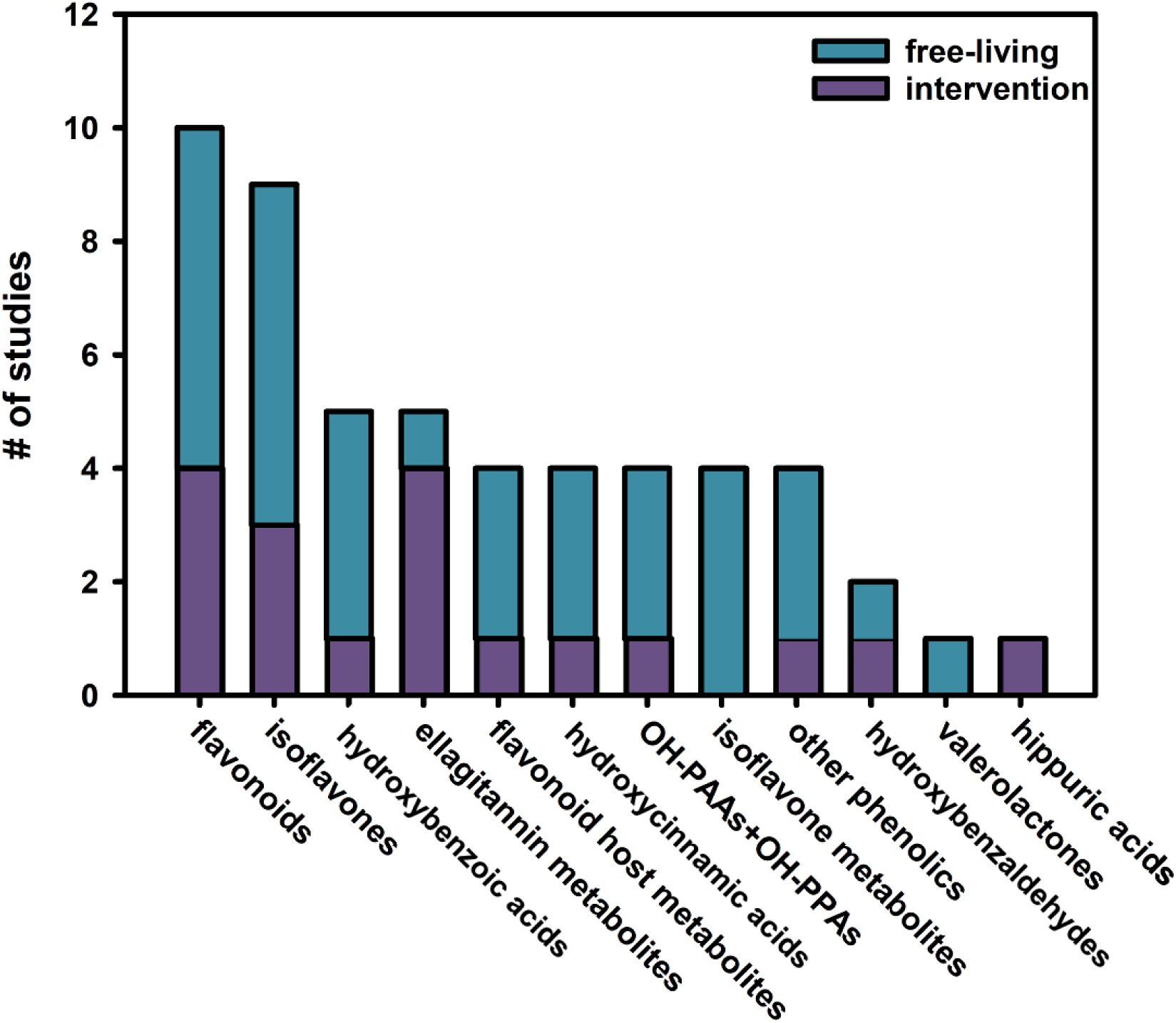
Total number of reports (n = 20) of polyphenols in human milk by separated by experimental design and subclass of polyphenols and metabolites. Reports come from single dose pharmacokinetic studies (n = 5), dietary interventions (2 d – 4 wks, n = 6), and observational cohorts (n = 10). OH-PAAs, hydroxyphenylacetic acids; OH-PPAs, hydroxyphenylpropanoic acids.

**Table 3.** Study characteristics for dietary interventions analyzing phenolic compounds in human milk.

| Study (Author, Year, Country) | Diet | Non-dietary factors | Duration | Milk Type | N | Dietary recommendations | Samples collected | Analysis of aglycones only? | Instrument | Compounds |
| --- | --- | --- | --- | --- | --- | --- | --- | --- | --- | --- |
| Cortés-Martín et al. (2020) [29] Spain | 30 g walnuts/d | urolithin metabotype | 3 d | mature | $\frac{11}{27}$ | avoid ellagitannin-containing foods | $\frac{24, 48, 72 \text{ hr}}{72 \text{ hr}}$ | No | HPLC-DAD-ESI-Q | urolithins |
| Franke et al. (1996) [42] USA | 5, 10, and 20 g soybeans/d | none | 0, 1, and 3 d, respectively | mature | 1 | none | pre-defined time points over 6 d | Yes | HPLC-DAD | isoflavones |
| Franke et al. (2006) [14] USA | 24 g soy protein beverage/d | none | 2-4 d | mature | 16 | avoid feeding soy-based infant formula | 1-2 hr after final soy beverage consumption | Yes | HPLC-DAD | isoflavones |
| Henning et al. (2019) [43] USA | 8 oz pomegranate juice/d | none | 2 wk | mature | 2 | low dietary polyphenol intake | day 1, 7, and 14 | No | LC-MS/MS | urolithins |
| Henning et al. (2022) [13] USA | 8 oz pomegranate juice/d | none | 2 wk | mature | 12 | low dietary polyphenol intake | day 0 and 14 | No | LC-MS/MS | urolithins |
| Jochum et al. (2017) [44] USA | 250 ml soy protein beverage/d | none | 6 d | mature | 18 | avoid flavonoid-rich foods and beverages | day 1, 4, 7, and 8 | Yes | HPLC | isoflavones, flavan-3-ols |
|  | 300 ml black tea/d |  |  |  | 18 |  |  |  |  |  |
|  | 30 ml water/d |  |  |  | 8 |  |  |  |  |  |
| Khymenets et al. (2016) [28] Spain | 40-50 g dark chocolate | none | single dose | mature | 2 | no other cocoa-containing products | pre-defined time points over 24 hr | No | UPLC-MS/MS | flavan-3-ol metabolites |
| Romaszko et al. (2017) [45] Poland | 250 ml red grapefruit juice | none | single dose | mature | 14 | avoid naringenin-containing products | pre-defined time points over 3 d | Yes | HPLC-DAD-MS | naringenin |
| Romaszko et al. (2014) [46] Poland | quercetin-rich onion soup | none | single dose | mature | 11 | avoid quercetin-containing products | pre-defined time points over 2 d | Yes | HPLC-PDA-ESI-MS | quercetin |
| Zhou et al. (2020) [47] China | 500 ml soy milk | none | single dose | NR | 6 | avoid isoflavone-containing products | pre-defined time points over 24 hr | Yes | UPLC-ESI-Qtrap | isoflavones |
| Fiecke et al. (2024) [48] USA | Mediterranean-style diet | FUT2 secretor phenotype | 4 wk | mature | 13 | none | day 0 and 28 | No | UPLC-MS/MS | polyphenols and phenolic metabolites |

NR, not reported; FUT2, α1,2-fucosyltransferase 2

**Table 4.** Study characteristics for cohort studies analyzing phenolic compounds in human milk in free-living lactating women.

| Study (Author, Year, Country) | Dietary or non-dietary factors | Cohort stratification<sup>a</sup> | Milk type | N | Analysis of aglycones only? | Instrument | Compounds |
| --- | --- | --- | --- | --- | --- | --- | --- |
| Khymenets et al. (2016) [28] Spain | dietary flavan-3-ol intake | none | C, T, M | 11 | No | UPLC-MS/MS | flavan-3-ol metabolites |
| Lu et al. (2021) [10] China | dietary polyphenol intake | 57.39-609.49 mg /d | M | 30 | Yes | UPLC-QQQ | phenolic compounds |
|  |  | 621.95-1,434.65 mg /d |  | 30 |  |  |  |
|  |  | 1,450.68-3,598.63 mg /d |  | 29 |  |  |  |
| Min et al. (2020) [49] China | none | none | NR | 50 | Yes | UPLC-QQQ | isoflavones |
| Nalewajko-Sieliwoniuk et al. (2020) [51] Poland | none | none | NR | 13 | Yes | UPLC-QQQ | phenolic compounds |
| Sánchez-Hernández et al. (2021) [30] Spain | breastfeeding duration | colostrum | C, T, M | 7 | No | UPLC-QQQ | phenolic compounds and metabolites |
|  |  | transitional |  | 7 |  |  |  |
|  |  | mature |  | 7 |  |  |  |
| Song et al. (2013) [27] USA | breastfeeding duration | wk 1 | T, M | 17 | Yes | HPLC-TOF-MS | flavonoids |
|  |  | wk 4 |  | 17 |  |  |  |
|  |  | wk 13 |  | 17 |  |  |  |
| Song et al. (2013) [52] Multinational | country (China, Mexico, USA), breastfeeding duration | wk 2 | T, M | 20 per country | Yes | HPLC-TOF-MS | flavonoids |
|  |  | wk 4 |  | 20 per country |  |  |  |
|  |  | wk 13 |  | 20 per country |  |  |  |
|  |  | wk 26 |  | 20 per country |  |  |  |
| Song et al. (2013) [52] Multinational | country (China, Mexico, USA), breastfeeding duration | wk 2 | T, M | 2 per country<sup>b</sup> | No | HPLC-TOF-MS | flavonoid metabolites |
|  |  | wk 4 |  | 2 per country<sup>b</sup> |  |  |  |
|  |  | wk 13 |  | 2 per country<sup>b</sup> |  |  |  |
|  |  | wk 26 |  | 2 per country<sup>b</sup> |  |  |  |
| Irvine et al. (1998) [53] New Zealand | dietary isoflavone intake | none | NR | 11 | Yes | HPLC | isoflavones |
| Setchell et al. (1998) USA | none | none | mature | 9 | Yes | GC-MS | isoflavones |
| Berger et al.<br>(2024) [54] Austria | breastfeeding duration | 1 mo | T, M | 2 | No | UHPLC-ESI-QTrap-MS/MS | phenolic<br>compounds and<br>metabolites |
|  |  | 6 mo |  | 2 |  |  |  |
|  |  | 12 mo |  | 2 |  |  |  |
| Berger et al.<br>(2024) [54] Austria | none | none | T, M | 30 | No | UHPLC-ESI-QTrap-MS/MS | phenolic<br>compounds and<br>metabolites |

<sup>a</sup>Factors (besides breastfeeding duration) used for cohort stratification.
<sup>b</sup>2 pooled samples derived from n = 20 per country were analyzed at each time point.
NR, not reported; C, colostrum; T, transitional; M, mature

##### Isoflavones and other flavonoids

Flavonoids, including isoflavones, are perhaps one of the most common classes of polyphenol detected and quantified in human milk from free-living lactating women. Common forms include flavan-3-ols, flavonols, flavanones, and isoflavones, with sparse reports of flavones and chalcones (**Table 5**) [10,27,30,50–54]. Individual flavan-3-ols include epicatechin (0.61-828.5 nmol/L), catechin (0.7-10.0 nmol/L), epicatechin gallate (0.08-645.6 nmol/L), and epigallocatechin gallate (2.54-1,683.8 nmol/L) [10,27,30,51,52,54]. Flavonols include quercetin (1.2-152.5 nmol/L), quercetin 3-*O*-glucoside (2.4-4.1 nmol/L), kaempferol (7.8-128.3 nmol/L), isorhamnetin (1.1 nmol/L) and kaempferol 3-*O*-glucoside (52.6-67.4 nmol/L) [10,27,30,52,54]. Flavanones include naringenin (0.03-722.0 nmol/L), 8-prenylnaringenin (3.82 nmol/L), and hesperetin (0.8-1,603.1 nmol/L) [10,27,51,52,54]. Isoflavones include daidzein (0.83-263.53 nmol/L) and genistein (0.3-117 nmol/L) [10,49–51,53,54]. The flavones apigenin (0.17-6.66 nmol/L) and diosmetin (<LOQ-5.33 nmol/L) have also been detected and quantified in human milk [54]. Only single reports exist analyzing human milk for epigallocatechin [27], quercetin 3-*O*-rutinoside [30], glycitein [49], proanthocyanidin C1 [54], and the chalcones phloretin and xanthohumol [54], which were not detected in any of these studies.

**Table 5.**
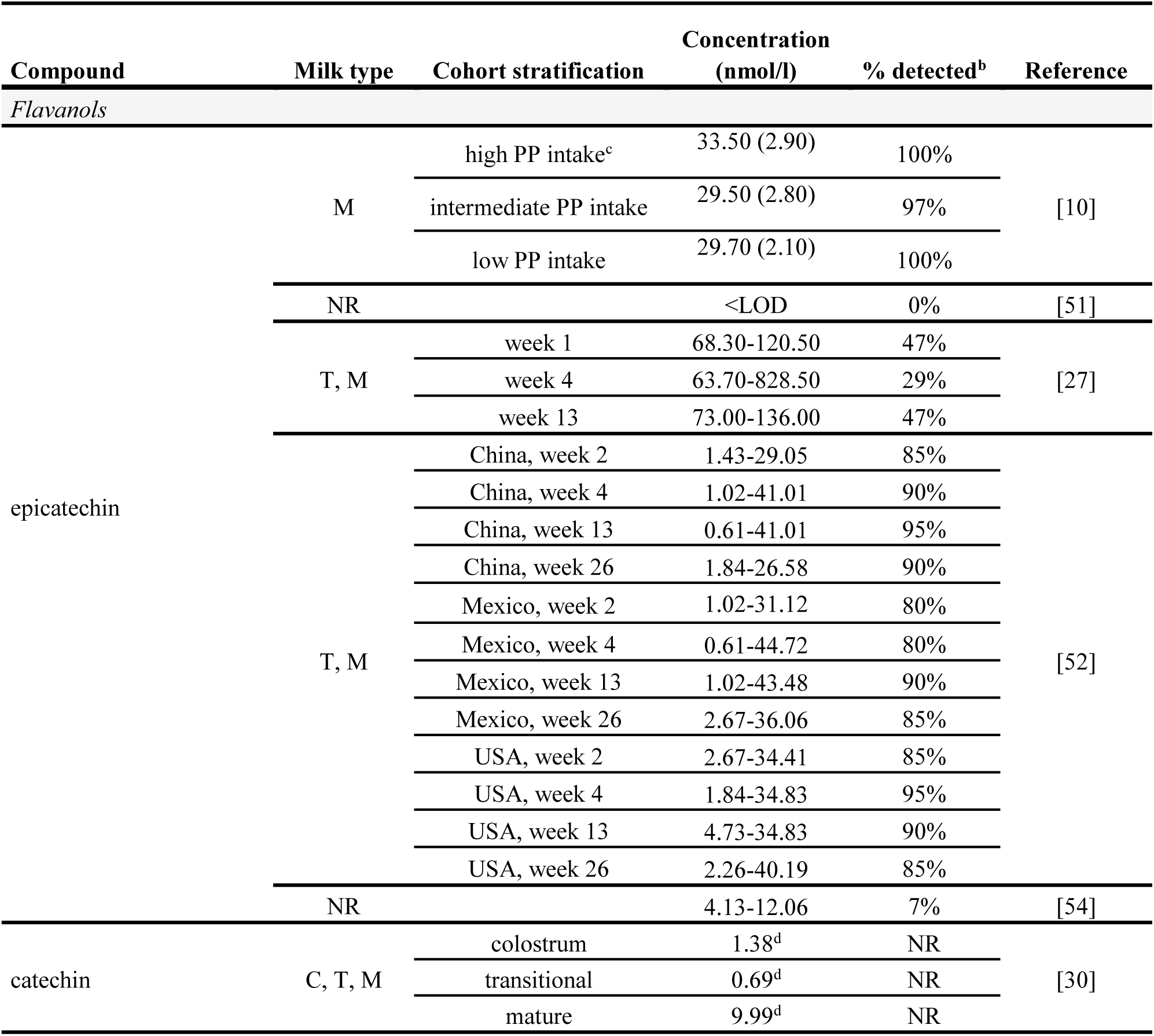
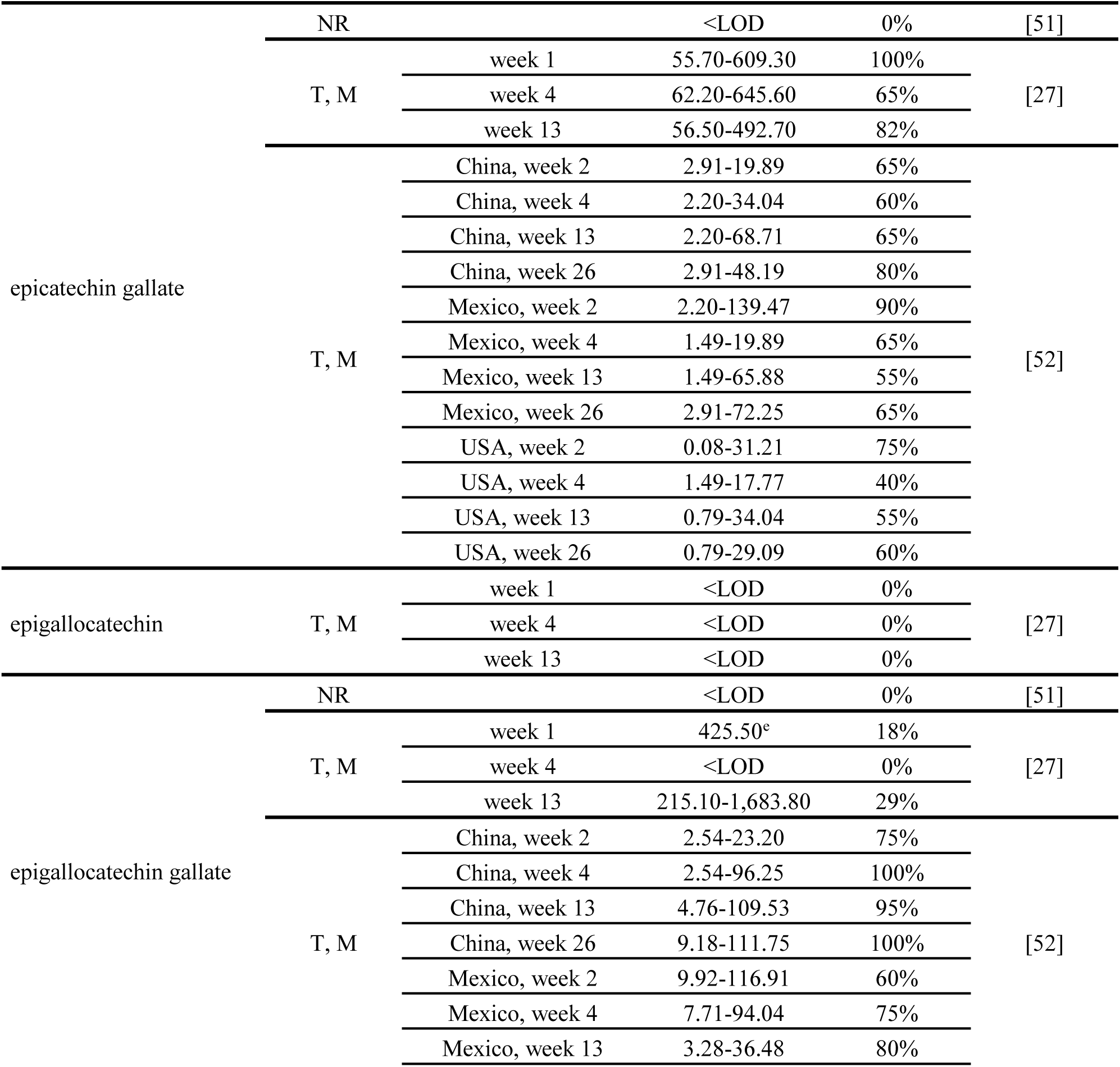
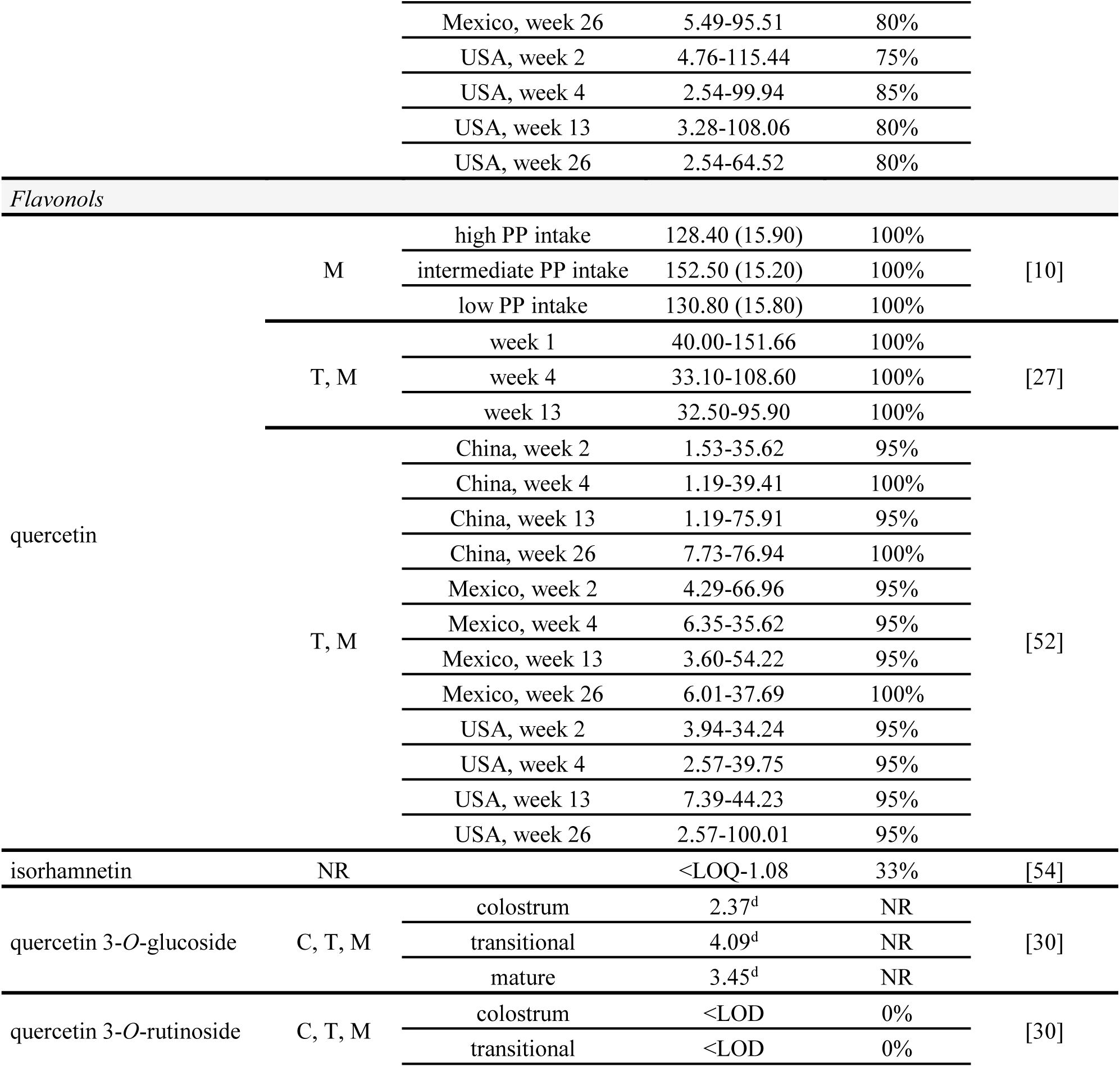
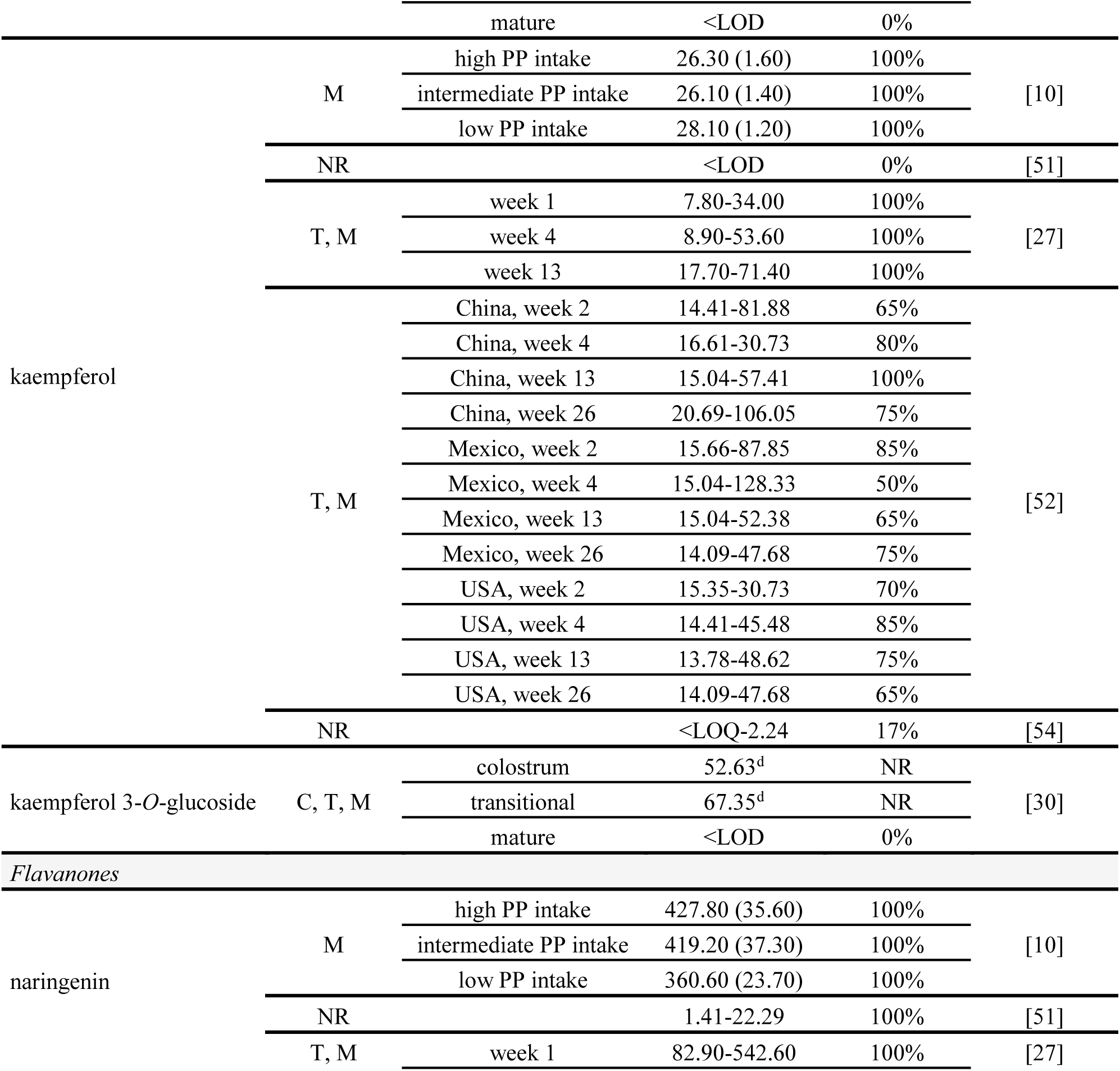
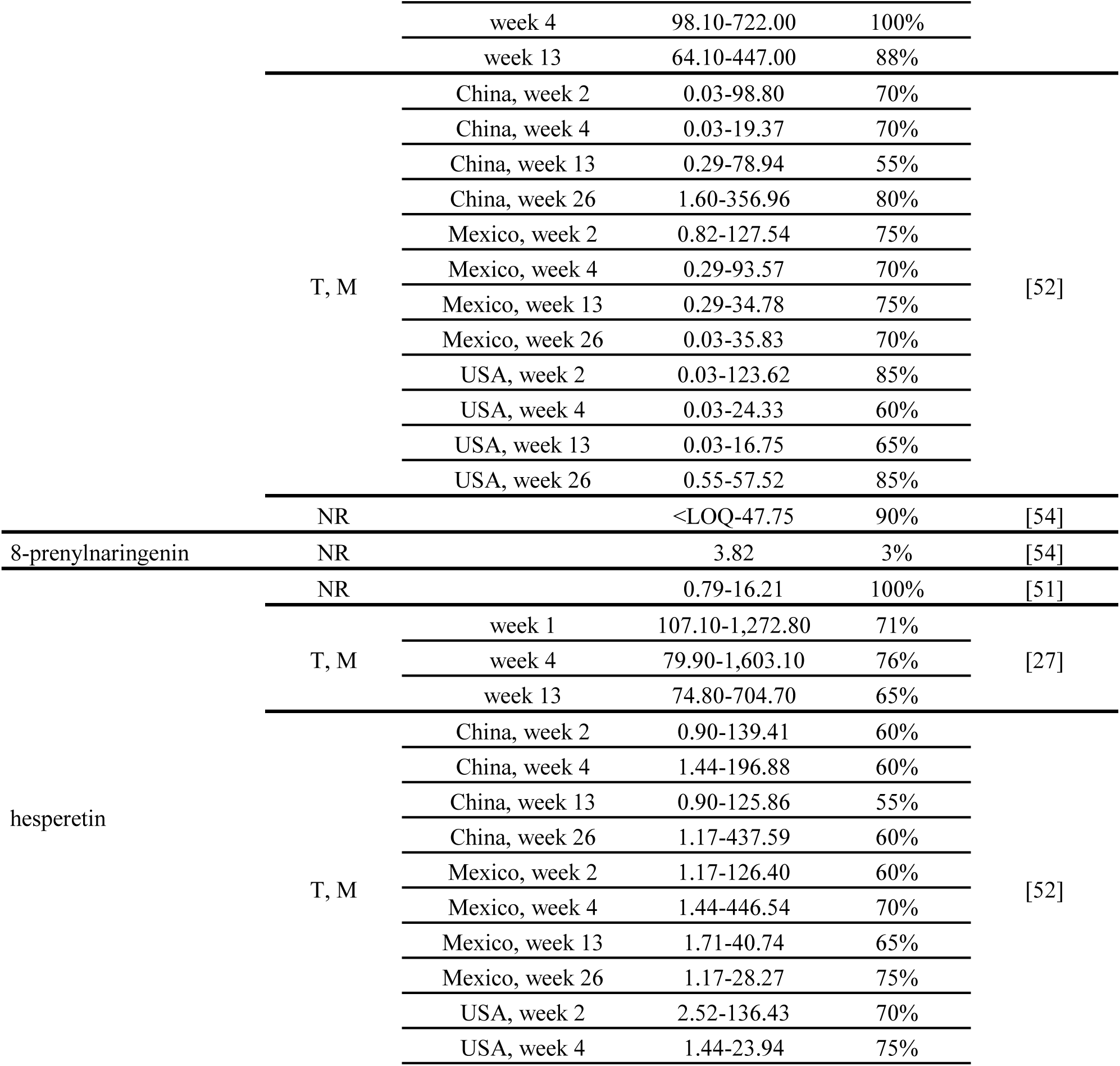
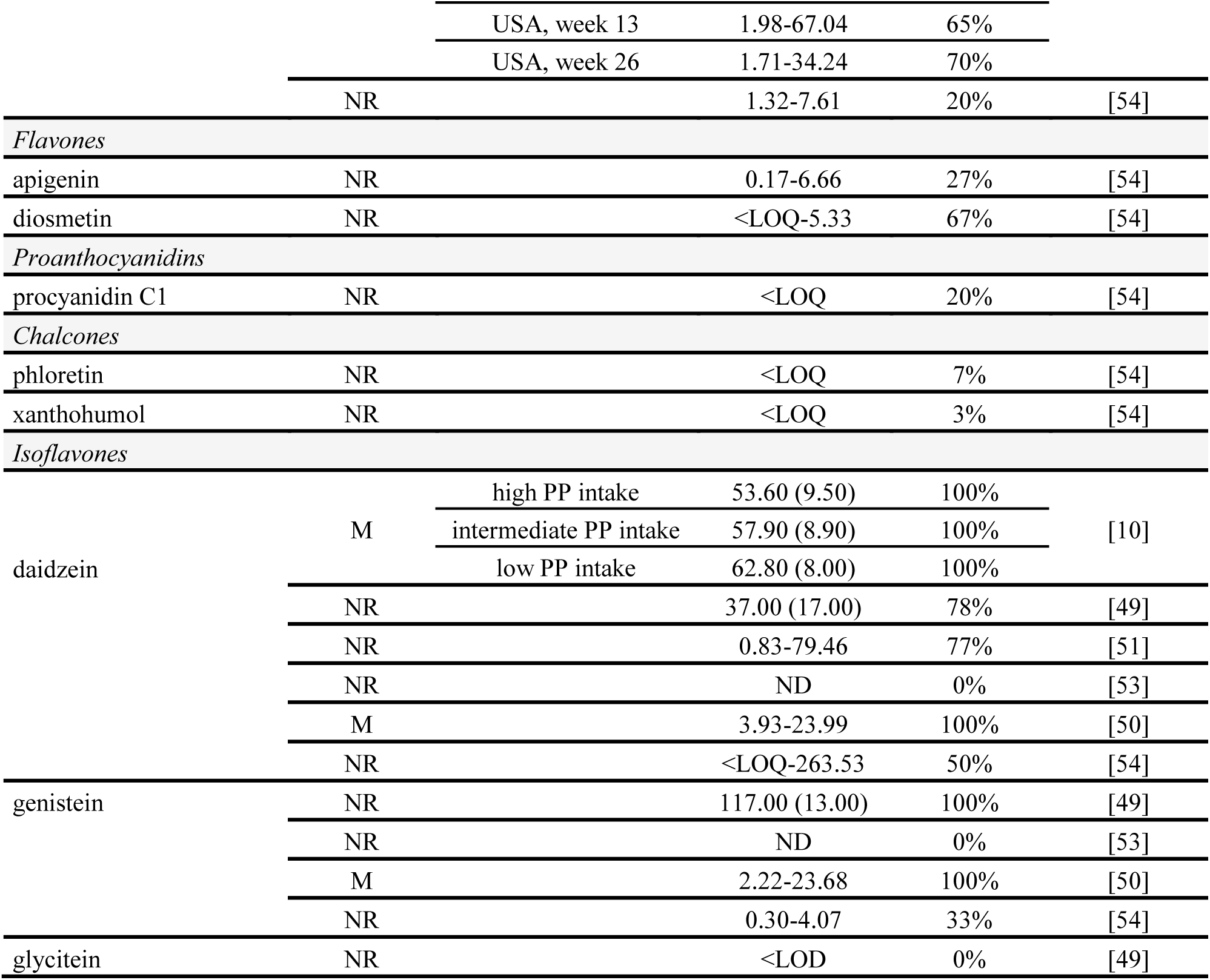
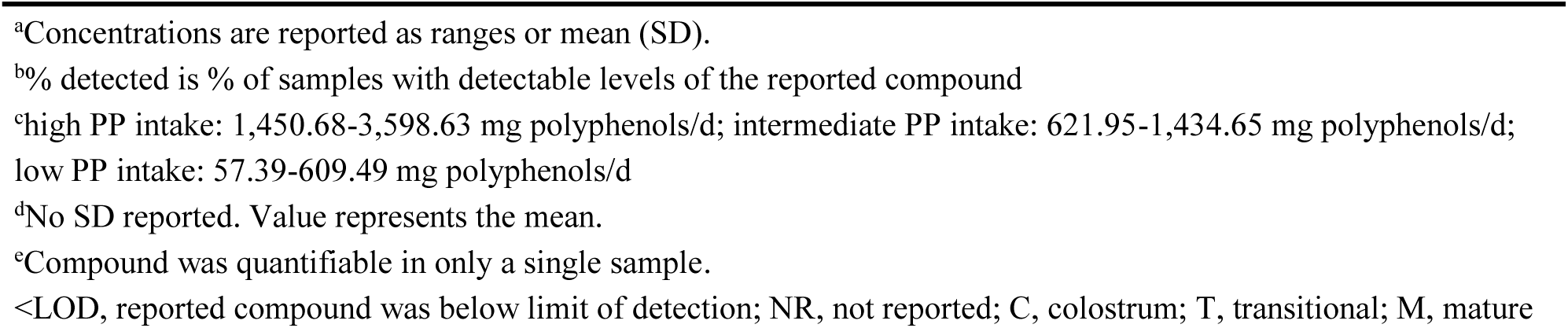
Flavonoids from food-derived sources detected and quantified in human milk in cohort studies in free-living lactating women^a^.

##### Flavonoid and isoflavone host metabolites

Methylated, glucuronidated, and sulfated metabolites of flavan-3-ols, flavonols, flavanones, and isoflavones have been detected and quantified in human milk by four studies (**Table 6**) [28,50,52,54]. Flavan-3-ol host metabolites include epicatechin glucuronide (0.04-36.4 nmol/L), epigallocatechin gallate glucuronide (5.0-8.5 nmol/L), epicatechin sulfate (13.4-14.5 nmol/L), methyl-epicatechin glucuronide (0.5-8.5 nmol/L), and methyl-epicatechin sulfate (13.2-23.7 nmol/L) [28,52]. Flavonol host metabolites include quercetin glucuronide (1.4-5.7 nmol/L), methyl-quercetin glucuronide (2.0-3.5 nmol/L), and kaempferol glucuronide (0.5-18.5 nmol/L) [52,54]. Flavanone host metabolites include naringenin glucuronide (0.1-0.6 nmol/L), methyl-naringenin glucuronide (1.5-10.4 nmol/L), and hesperetin glucuronide (0.1-4.8 nmol/L) [52]. Isoflavone host metabolites include daidzein 7-*O*-glucuronide (0.3-1.37 nmol/L), genistein 7-*O*-glucuronide (<LOQ-4.3 nmol/L), and genistein 7-*O*-sulfate (<LOQ) [54]. Certain metabolites, including sulfated epicatechin metabolites, have been detected in a single study, with Khymenets et al. [28] reporting that only 3/24 (13%) and 7/24 (29%) of samples contained detectable concentrations of epicatechin sulfate and methyl-epicatechin sulfate, respectively.

**Table 6.**
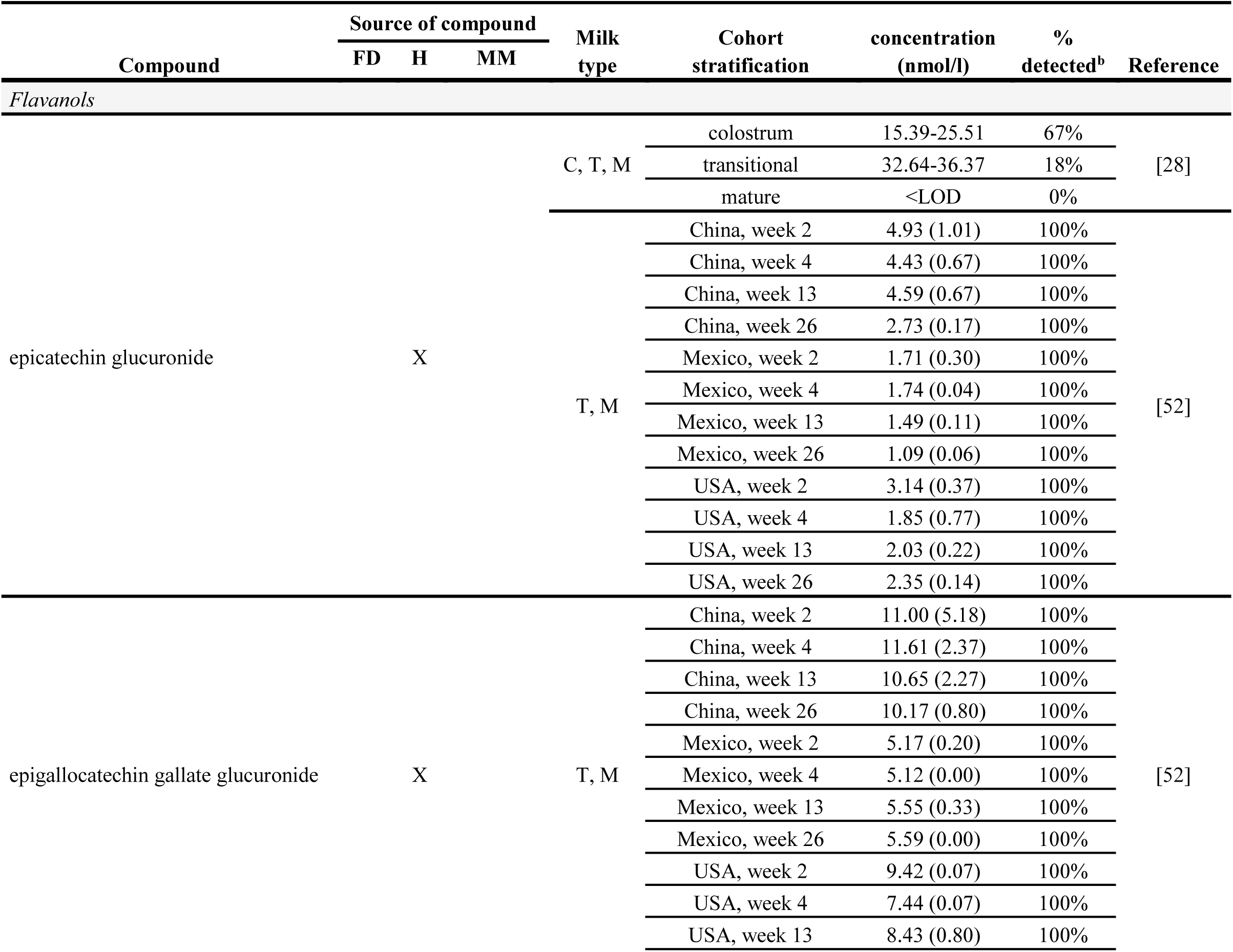
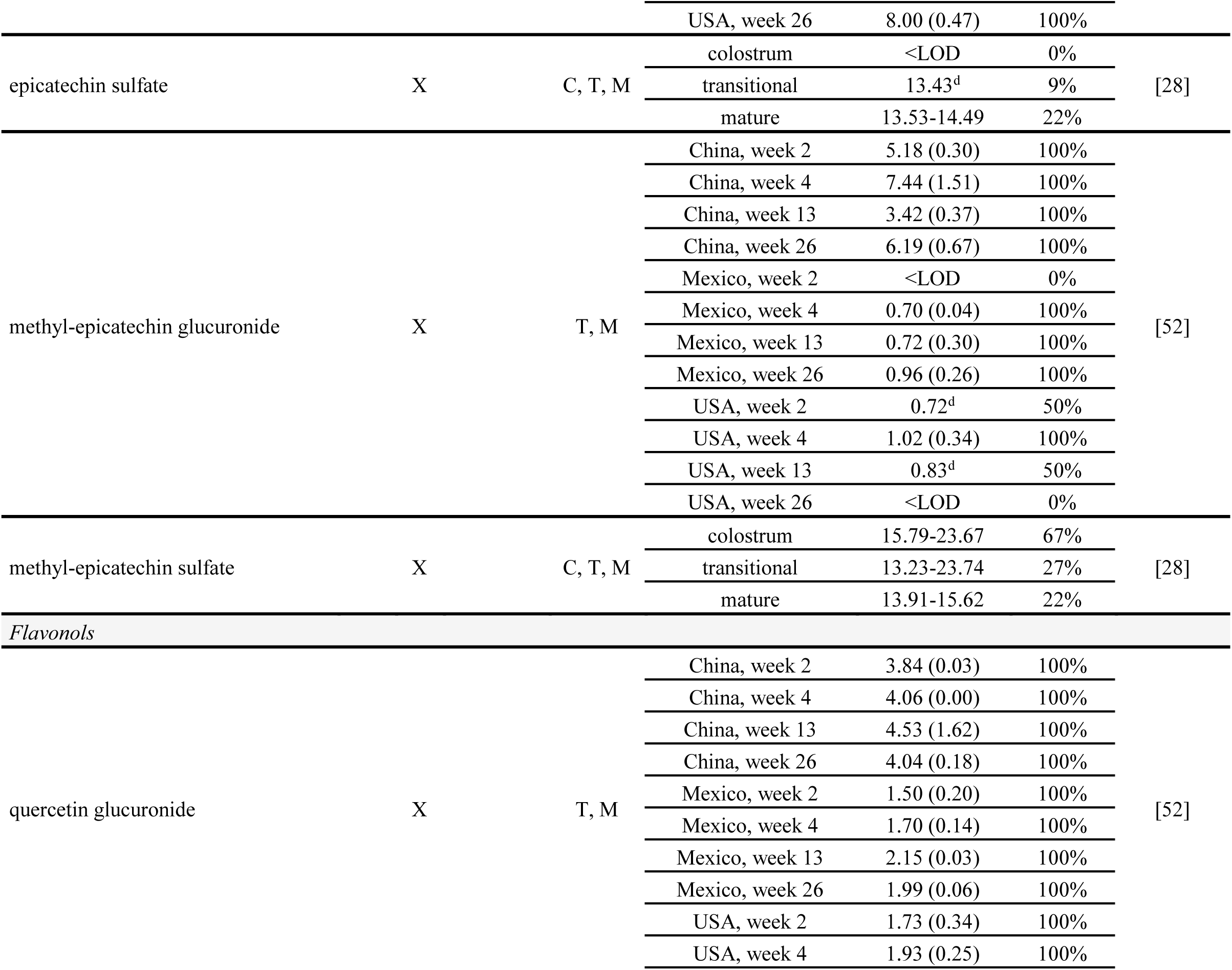
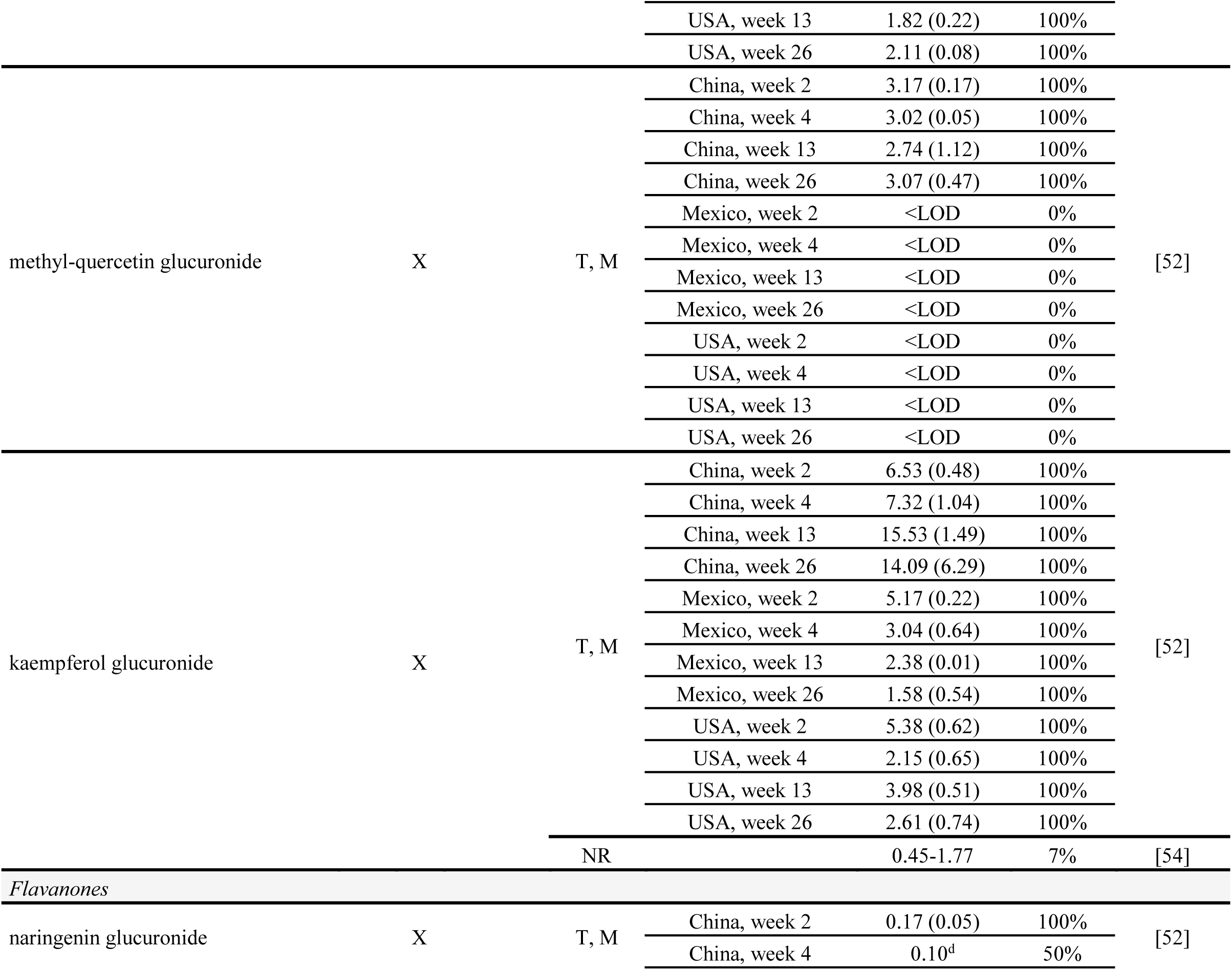
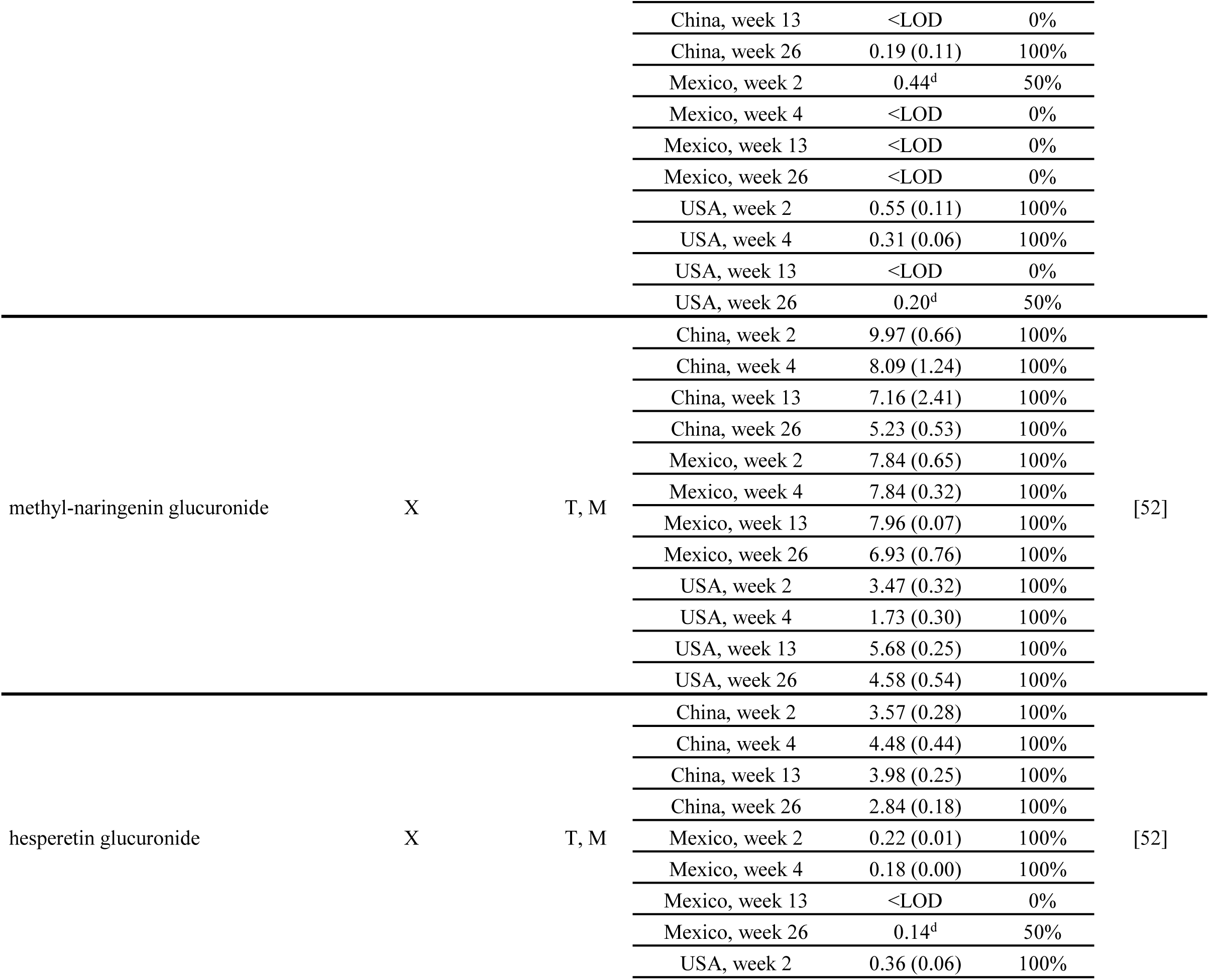
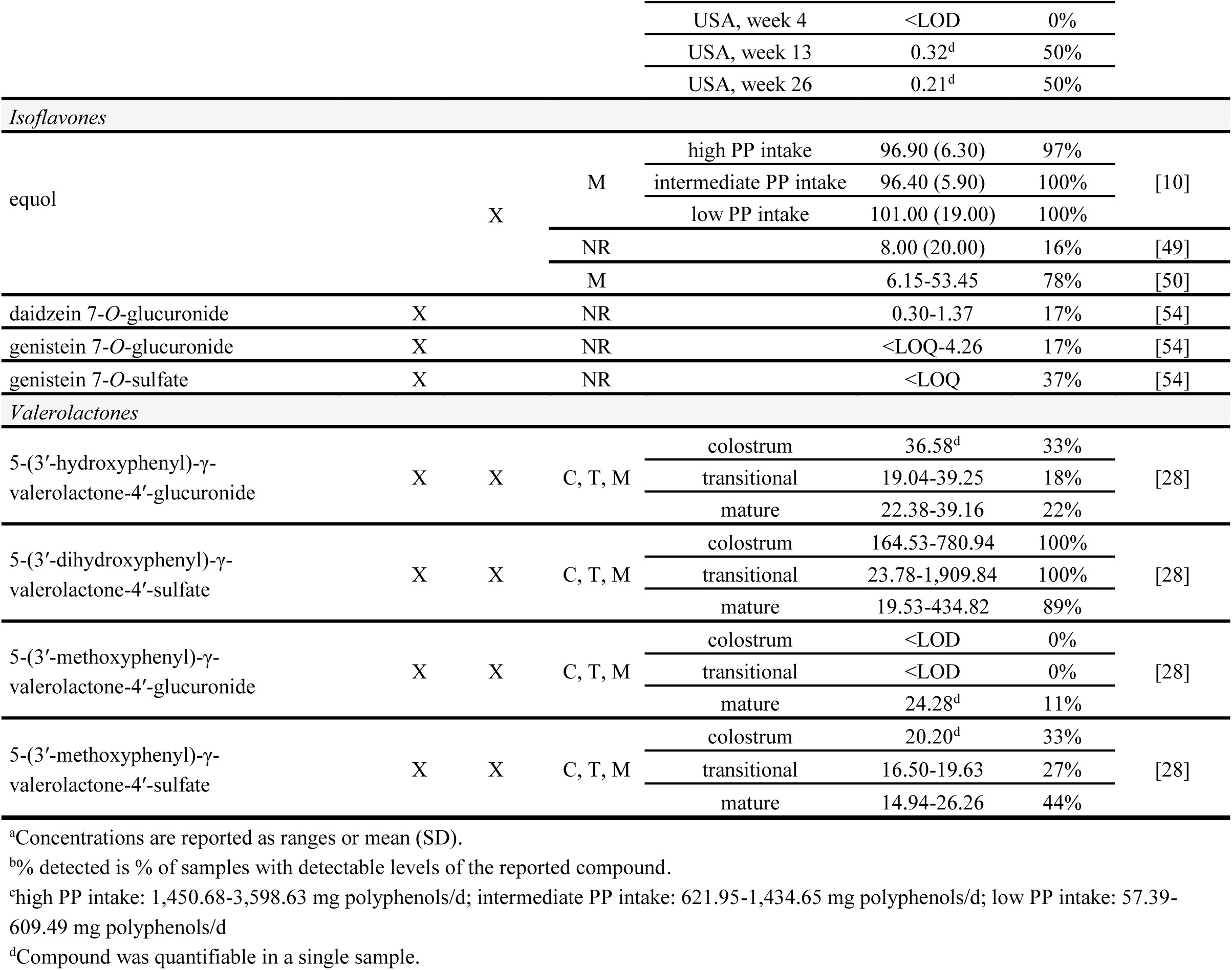
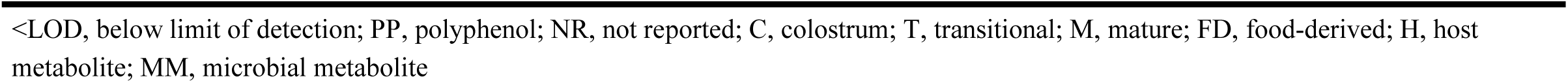
Flavonoid metabolites detected and quantified in human milk in cohort studies in free-living lactating women^a^.

##### Phenolic acids and other microbial-derived metabolites

Various phenolic acids derived from food or through microbial metabolism of dietary polyphenols have been quantified in human milk, in addition to other microbial-derived polyphenol metabolites. These include hydroxybenzoic acids [10,30,51,54], hydroxycinnamic acids [10,30,54], hydroxybenzaldehydes [30], hydroxyphenylacetic acids [10,30,54], hydroxyphenylpropanoic acids [54], valerolactones [28], equol [10,49,50], enterodiol [54], enterolactone [49,54], and urolithins [54] (**Table 6** and **Table 7**). Hydroxybenzoic acids include: 3,4,5-trihydroxybenzoic acid (0.6-194.6 nmol/L); 3,4-dihydroxybenzoic acid (851.9-1,463.2 nmol/L); 3,5-dimethoxy-4-hydroxybenzoic acid (49.5 nmol/L); 4-hydroxy-3-methoxybenzoic acid (2.6-419.3 nmol/L); 3,5-dihydroxybenzoic acid (2,595.4 nmol/L); 2,4,6-trihydroxybenzoic acid (157.5-181.1 nmol/L); 2,5-dihydroxybenzoic acid (364.0-480.8 nmol/L); 2-hydroxybenzoic acid (10.1-2,606.4 nmol/L); 3-hydroxybenzoic acid (311.3-719.7 nmol/L); 4-hydroxybenzoic acid (39.1-2,968.4 nmol/L); and ethyl gallate (0.3 nmol/L) [10,30,51,54]. Hydroxycinnamic acids include: 4-methoxycinnamic acid (13.5-51.1 nmol/L); cinnamic aicd (108.0-162.0 nmol/L); 3ʹ,4ʹ-dimethoxycinnamic acid (8.7-26.9 nmol/L); 3-*O*-caffeoylquinic acid (5.0-500.1 nmol/L); 4ʹ-hydroxy-3ʹ-methoxycinnamic acid (17.5-119.4 nmol/L); 3ʹ,4ʹ-dihydroxycinnamic acid (15.0-959.2 nmol/L); 4ʹ-hydroxycinnamic acid (17.7-140.1 nmol/L); and 4ʹ-hydroxy-3ʹ,5ʹ-dimethoxycinnamic acid (8.92 nmol/L) [10,30,54]. Only one study has detected and quantified hydroxybenzaldehydes in human milk from free-living lactating women. Sánchez-Hernández et al. [30] quantified the hydroxybenzaldehydes 2- hydroxy-3-methoxybenzaldehyde (105.8-803.2 nmol/L), 4-hydroxy-3-methoxybenzaldehyde (365.4-514.6 nmol/L), 4-hydroxybenzaldehyde (132.7-289.9 nmol/L), and 3,4- dihydroxybenzaldehyde (540.1-909.4 nmol/L). Hydroxyphenylacetic acids include 3ʹ,4ʹ- dihydroxyphenylacetic acid (26.2-386.6 nmol/L) and 4ʹ-hydroxyphenylacetic acid (17.8-78,869.5 nmol/L) [10,30,54]. Hydroxyphenylpropanoic acids have only been detected and quantified by Berger et al. [54], demonstrating the presence of 3-(3ʹ-hydroxyphenyl)propanoic acid (463.4 nmol/L), 3-(3ʹ,4ʹ-dihydroxyphenyl)propanoic acid (1,591.9 nmol/L), 3-(4ʹ-hydroxy-3ʹ- methoxyphenyl)propanoic acid (6.1-42.3 nmol/L) in human milk.

**Table 7.**
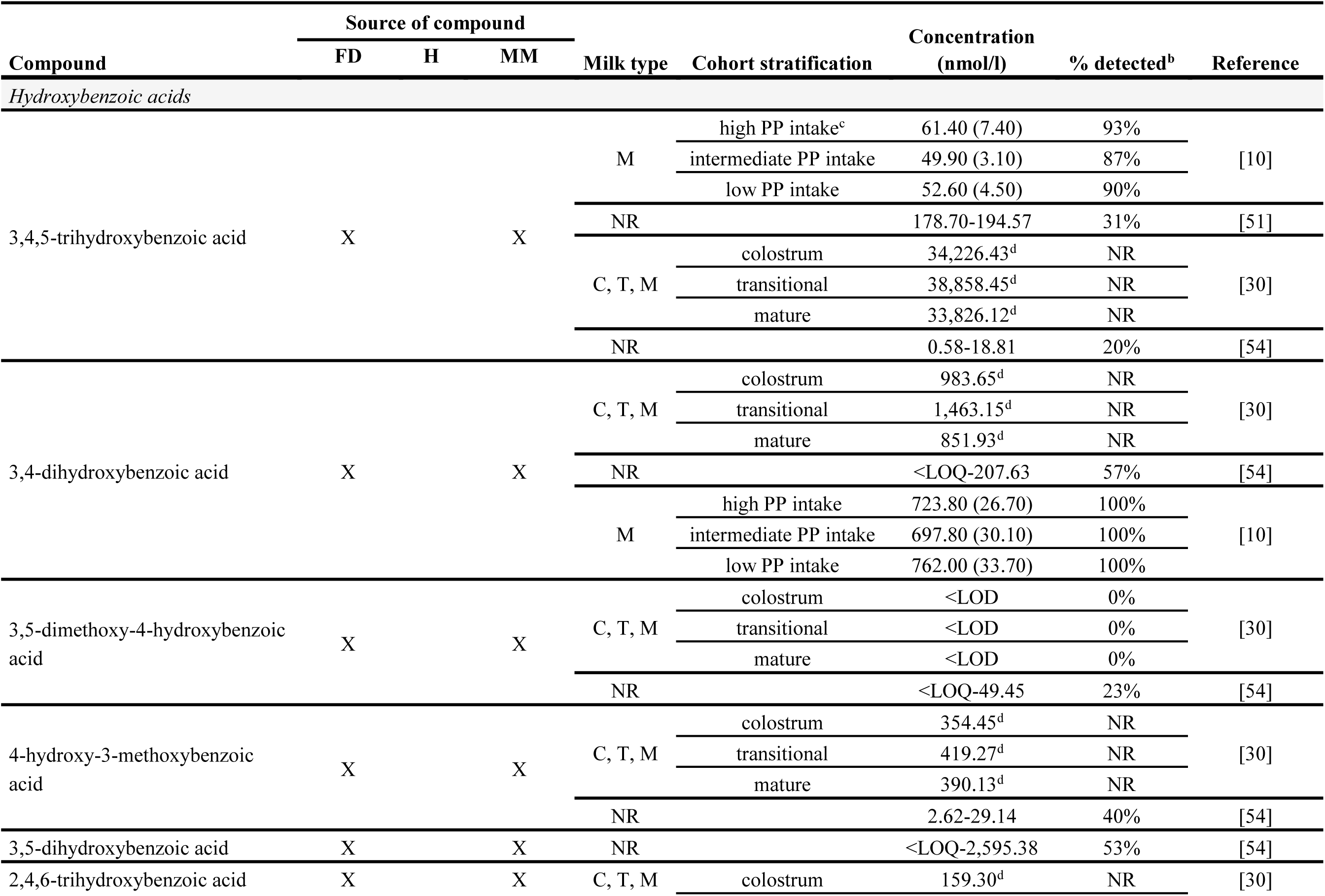
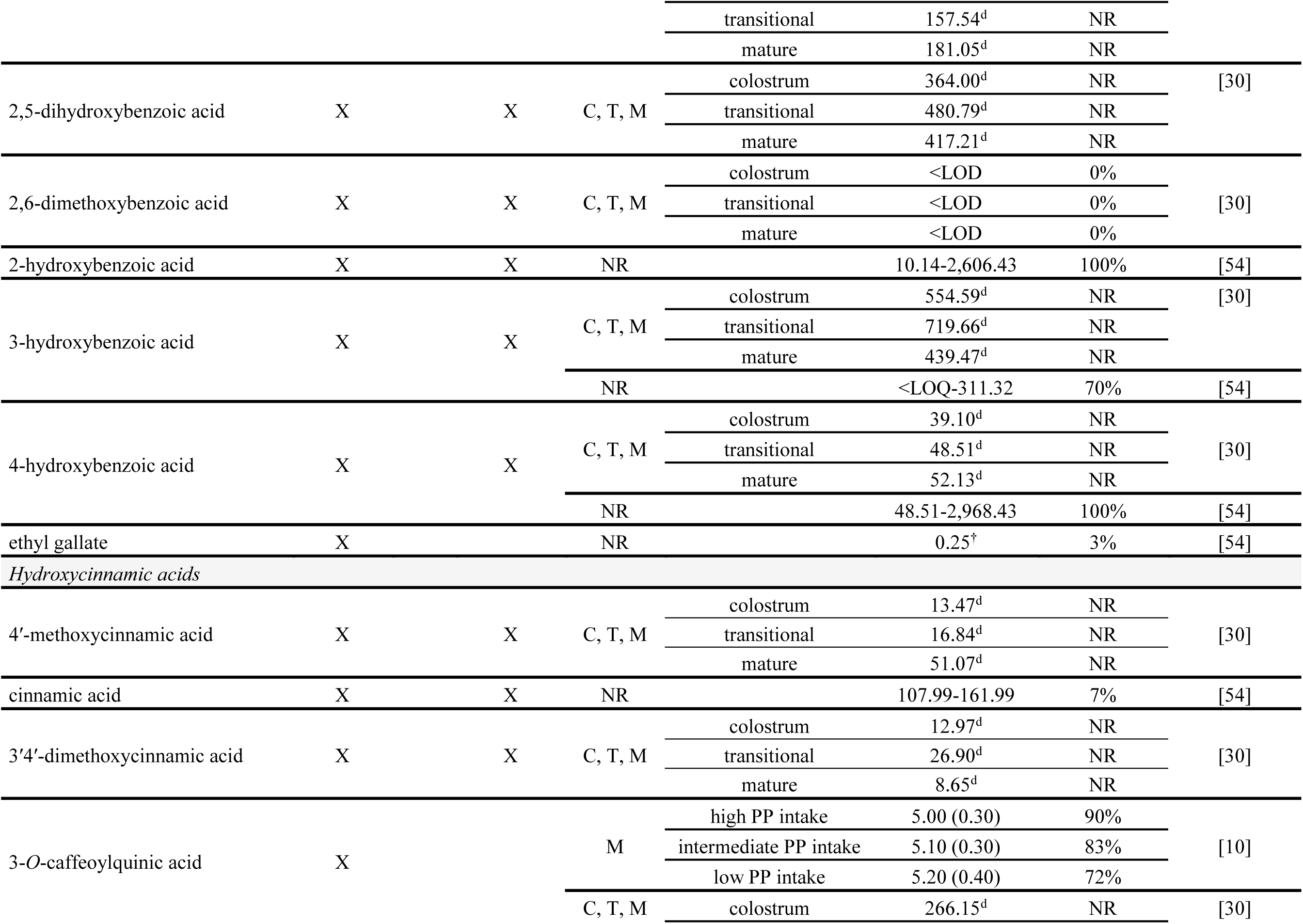
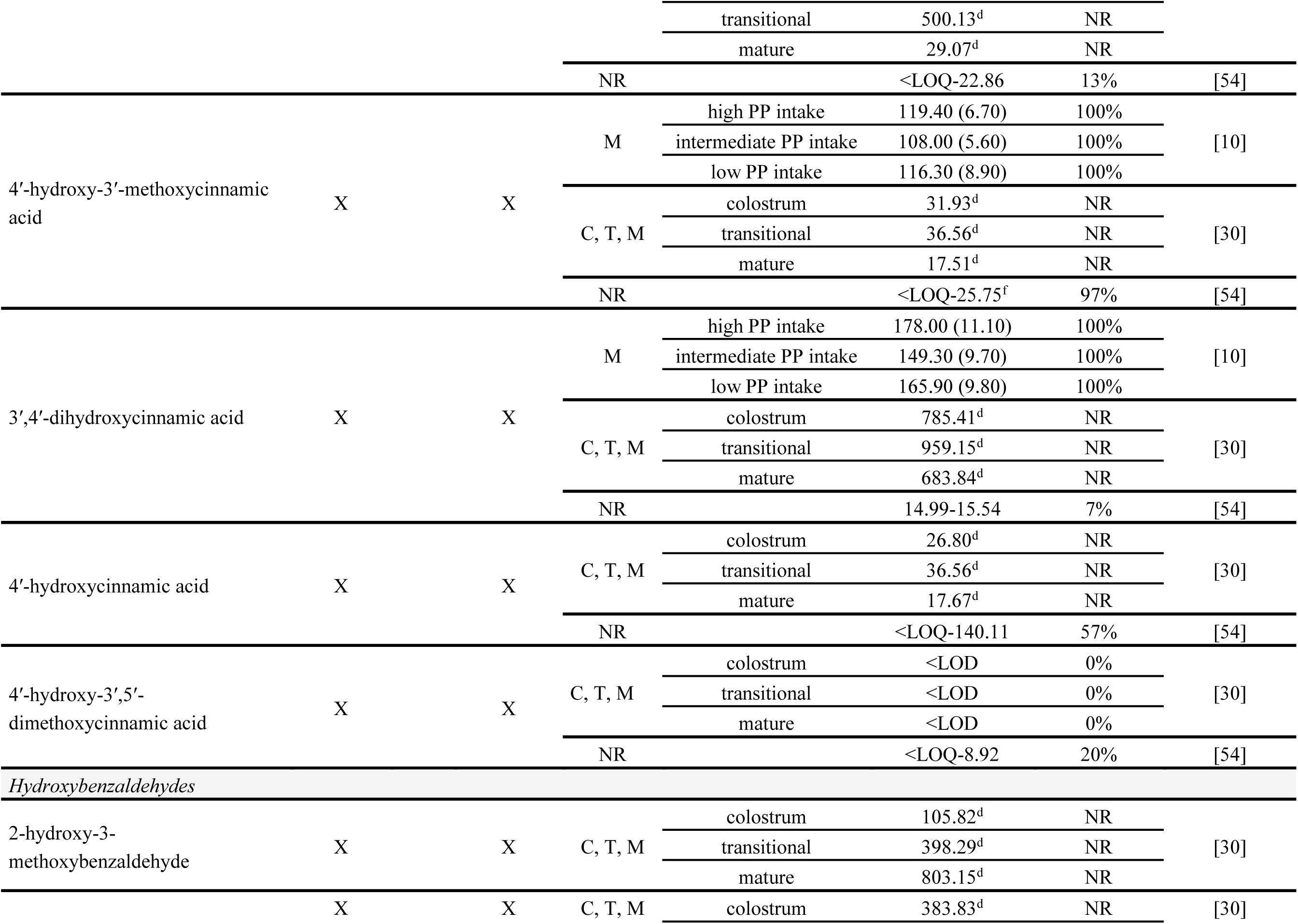
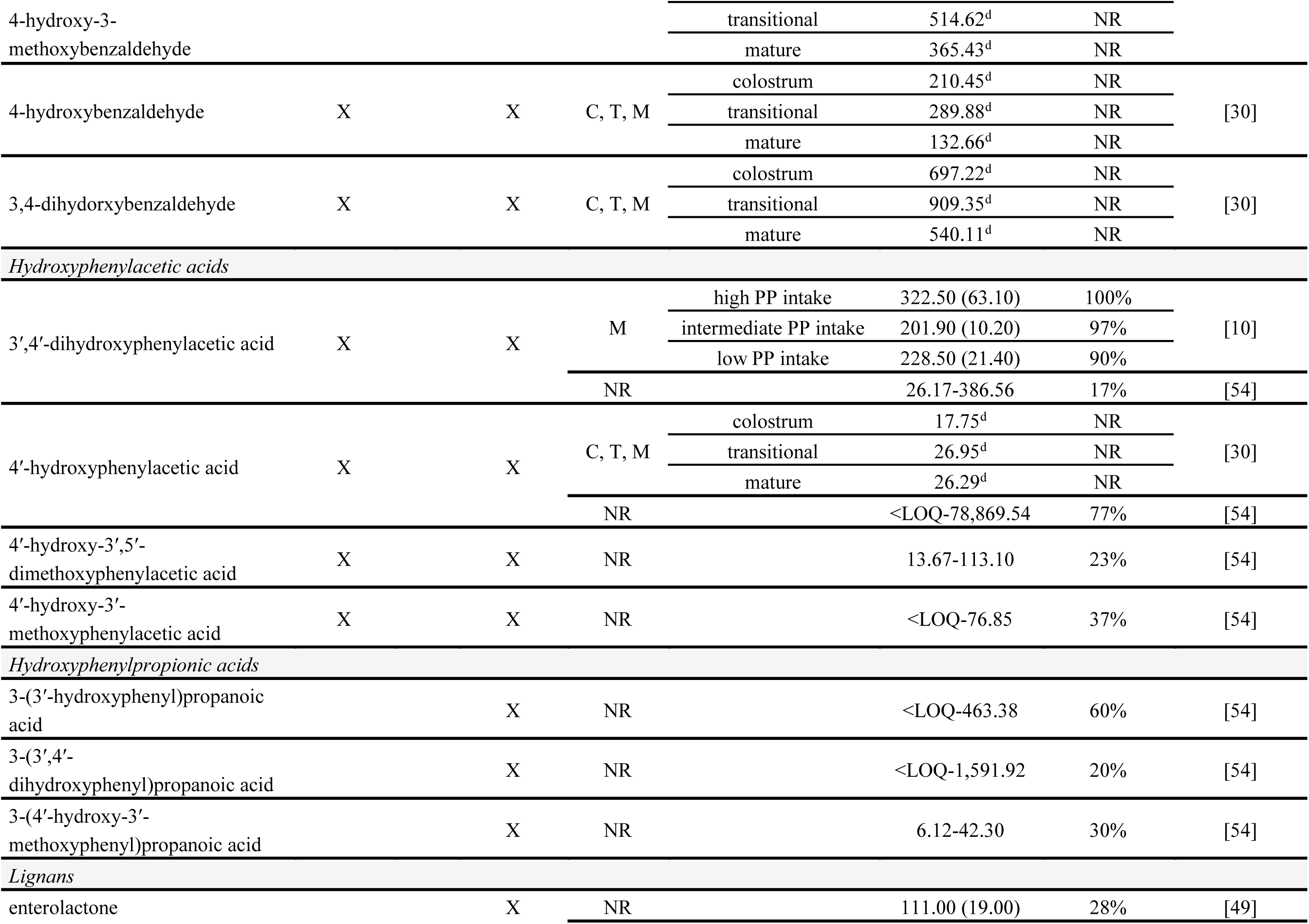
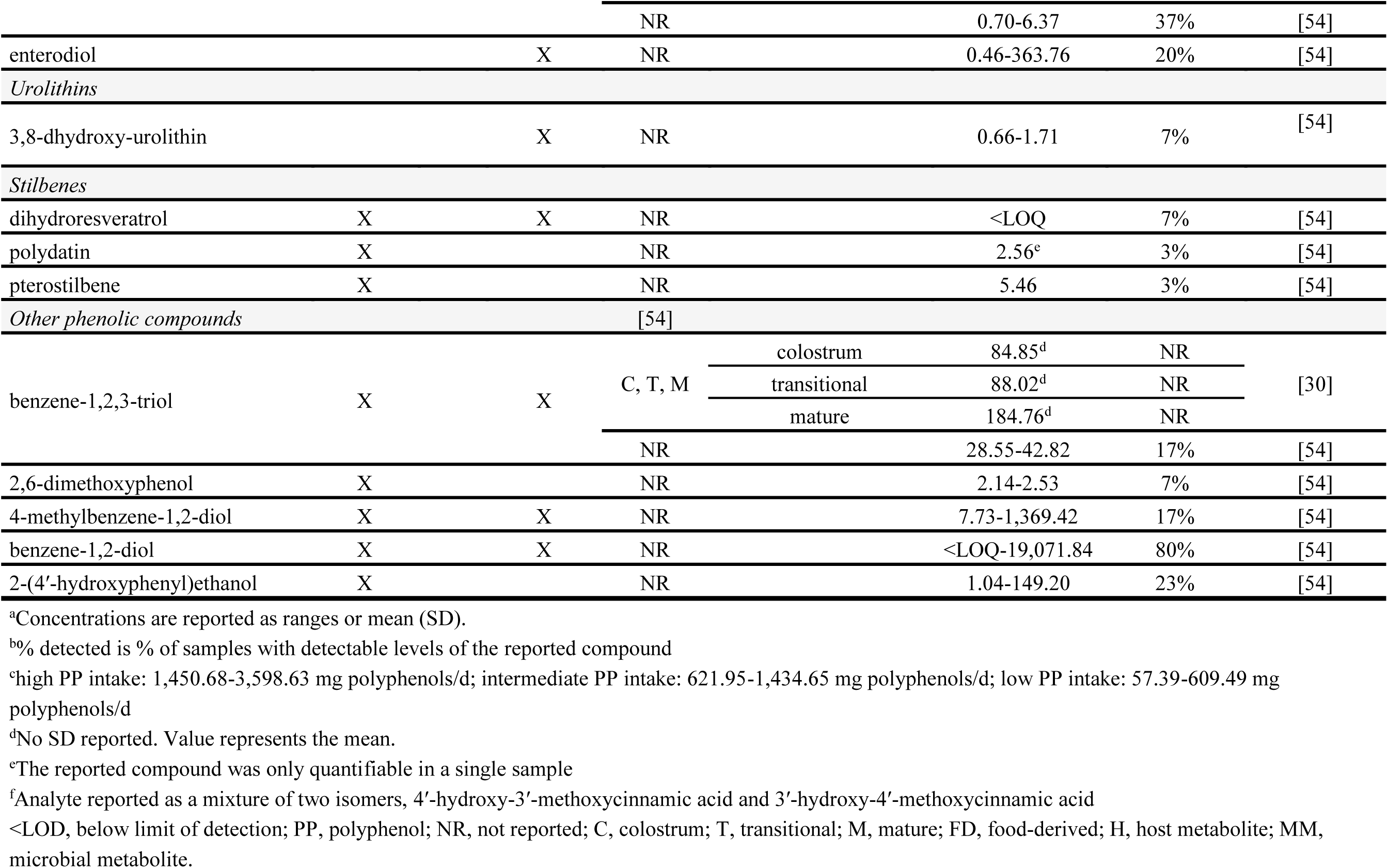
Phenolic acids, hydroxybenzaldehydes, and other phenolic compounds detected and quantified in human milk in cohort studies in free-living lactating women.^a^.

Valerolactones are metabolites derived from gut microbial metabolism of flavan-3-ols that have only been detected and quantified by Khymenets et al [28]. They quantified 5-(3ʹ- dihydroxyphenyl)-γ-valerolactone-4ʹ-glucuronide, 5-(3ʹ-dihydroxyphenyl)-γ-valerolactone-4ʹ- sulfate, 5-(3ʹ-methoxyphenyl)-γ-valerolactone-4ʹ-glucuronide, and 5-(3ʹ-methoxyphenyl)-γ- valerolactone-4ʹ-sulfate in human milk. Of these metabolites, 5-(3ʹ-dihydroxyphenyl)-γ- valerolactone-4ʹ-sulfate was detected and quantified most consistently (>90% of samples) compared to the others (<25% of samples). 5-(3ʹ-methoxyphenyl)-γ-valerolactone-4ʹ-glucuronide was detected and quantified least frequently (4% of samples). Average concentrations ranged between 19 and 30 nmol/L, with 5-(3ʹ-dihydroxyphenyl)-γ-valerolactone-4ʹ-sulfate demonstrating the largest range (19.5-8,305 nmol/L).

Equol is a gut microbial metabolite of isoflavones [49] that has been measured in three studies, with concentrations in the range of 6.2-101.0 nmol/L [10,49,50]. Although Min et al. reported a low proportion of samples in which equol was detected (<20% of samples) [49], other reports are generally more consistent (>75% of samples) [10,50].

Enterolactone and enterodiol are gut microbial metabolites of lignans [90] and have been quantified in two studies, demonstrating concentrations in the range of 0.7-111.0 nmol/L and 0.5-363.8 nmol/L, respectively [49,54]. However, the proportion of samples in which these lignan metabolites have been detected tends to be lower (<40%) [49,54].

Urolithins have been quantified in one free-living cohort, demonstrating low, sparse concentrations of 3,8-dihydroxy-urolithin (0.7-1.7 nmol/L; detected in 7% of samples), a microbial metabolite of ellagitannins [54]. Although most phenolic acids are consistently reported in human milk, sparse detection of 3,5-dimethoxy-4-hydroxybenzoic acid (detected in <25% of samples), ethyl gallate (<5% of samples), and 4ʹ-hydroxy-3ʹ,5ʹ-dimethoxycinnamic acid (<25% of samples) have been reported by Sánchez-Hernández et al. and Berger et al. [30,54]. However, additional studies are needed to confirm these findings due to the paucity of available data.

##### Stilbenes and other phenolic compounds

There are only two studies that have reported concentrations of stilbenes, phenols, and tyrosols in observational cohorts [30,54]. Stilbene concentrations in human milk tend to be low (<10 nmol/L; detected in <10% of samples), with only a single report detecting dihydroresveratrol, polydatin, and pterostilbene [54]. Phenols and tyrosols that have been detected and quantified in human milk include: benzene-1,2,3-triol (28.6- 184.8 nmol/L); 2,6-dimethoxyphenol (2.1-2.5 nmol/L); 4-methylbenzene-1,2-diol (7.7-1,369.4 nmol/L); benzene-1,2-diol (19,071.8 nmol/L); and 2-(3ʹ-hydroxyphenyl)ethanol (1.0-149.2 nmol/L) [30,54].

#### 3.1.3. Transfer of polyphenolic compounds and metabolites from human milk to circulation of breastfed infants

A limited number of studies [13,14,49] have directly demonstrated that polyphenols and their metabolites can be transferred from human milk to infant circulation through breastfeeding. The urine of breastfed infants has been shown to contain quantifiable concentrations of isoflavones derived from maternal soy intake [14] and urolithins derived from maternal dietary intake of ellagitannins (e.g., pomegranate juice) [13]. Following a 15 d dietary intervention of 8 oz/d pomegranate juice in healthy, lactating women, human milk was found to contain 8-hydroxy-urolithin-3-glucuronide (3.2-28.7 nmol/L) and urolithin-3-glucuronide (0.5-11.9 nmol/L) [13].

These urolithins were subsequently transferred to the breastfed infant, with urinary concentrations reaching 8.9-40.3 nmol/L and 0-17.5 nmol/L, respectively. It has also been shown that total isoflavone concentrations in human milk and urine of breastfed infants increase by 14-fold and 4-fold, respectively, after 2-4 d maternal soy intake (∼55 mg/d total isoflavones) [14].

The differing fold changes between human milk and infant urinary excretion may be related to infant milk intakes, which were not quantified. Polyphenol metabolites derived from maternal dietary intake can be transferred from human milk to infant circulation through breastfeeding. Polyphenols that are bioavailable to breastfed infants from human milk include urolithins (derived from maternal pomegranate juice consumption) [13] and isoflavones (derived from maternal soy consumption) [14], which have been quantified in the urine of breastfed infants.

Historically, interest in the transfer of soy isoflavones to infants has been driven by the prevalence of soy-based infant formulas. This has resulted in studies exploring the presence of soy isoflavones in human milk [10,49–51,53,54], as well as infant formulas [53], but still few reports exist directly linking maternal soy intake to transfer of soy isoflavones or their metabolites in breastfed infants [14,49]. Isoflavones have been simultaneously quantified in human milk and in the urine of breastfed infants in one observational cohort [49]. Although only genistein was quantifiable in human milk (117.0 ± 13.0 nmol/L), genistein (96.0 ± 16.0 nmol/L; detected in 50/50 samples), daidzein (44.0 ± 95.0 nmol/L; detected in 36/50 samples), and glycitein (232 ± 354 nmol/L; detected in 16/50 samples) were quantifiable in the urine of breastfed infants, suggesting maternal-infant transfer from human milk. Urinary excretion of the microbial isoflavone metabolite equol (47.0 ± 104.0 nmol/L) was also observed for 33/50 of the breastfed infants. Existing evidence suggests that there is a relationship between maternal dietary intake and concentrations of polyphenols and their metabolites in human milk and urine of breastfed infants, which is indicative of these compounds transferring from maternal diet to human milk to breastfed infants. However, studies reporting concentrations of polyphenols in human milk have not yet simultaneously evaluated volume of human milk intake to quantify polyphenol intakes by breastfed infants. As such, the rate and extent of transfer and exposure to these compounds from maternal diets to breastfed infants remains to be explored in detail.

### 3.2. Factors influencing concentrations of polyphenolic compounds and metabolites in human milk

#### 3.2.1. Potential factors influencing polyphenol composition of human milk

Maternal factors (e.g., genetics, diet, health status) have been evaluated in relation to composition of polyphenols or phenolic metabolites in human milk, although this has been reported only in a limited number of studies. Dietary polyphenols require absorption in the intestines and subsequent transfer from the circulation to human milk. Since the majority of polyphenols in circulation [91] and human milk [28,30,54] are small microbial metabolites, transfer to human milk from circulation likely occurs by passive diffusion. Factors known to influence absorption, distribution, metabolism, and excretion of dietary polyphenols include acute and chronic dietary intake [31,92], dose [93], food matrix factors [74,94–96], type of dietary polyphenols consumed [74,75,97], physical activity [98], health status [92,99], host genetics [100], and gut microbiome composition [101] are likely to have an influence on circulating profiles of polyphenols and their metabolites. The relationship between timing of maternal dietary intake and human milk collection is also likely to impact profiles and concentrations of milk polyphenols. However, a limited number of factors have been explored as potential factors influencing human milk, including lactation duration [27,28,30,52], maternal demographics [27,52], metabotype [29], genetics [48], and dietary polyphenol intake [13,14,29,42–48]. Other factors known to influence composition of other human milk components but that have not yet been explored in the context of polyphenol profiles, include diurnal fluctuations [102] and compositional changes over the course of a feed [103].

#### 3.2.2. Lactation stage and polyphenol profiles of human milk

Few longitudinal studies of polyphenol profiles/mixtures in human milk have been carried out, most of which included only flavonoids or flavonoid metabolites [27,28,52], with only one report of a broader range of polyphenols and phenolic metabolites [28]. Song et al. reported concentrations of various flavonoids (flavan-3-ols, flavonols, flavanones) across different lactation stages (weeks 1, 4, and 13 postpartum) in a subset of the Global Exploration of Human Milk study [104] that included 17 women from a U.S. cohort of lactating women [27]. In this cohort, a significant increase (∼1.2-fold) in kaempferol concentration was observed in week 13. These data were expanded to include a multinational cohort of women from China, Mexico, and the U.S. [52]. Significant associations with lactation stage were observed for epicatechin gallate in the cohort from Mexico and for flavan-3-ols and flavonols in the cohort from China (**Fig. 4**). Because both of these analyses [27,52] only evaluated associations with lactation stage [27], it is possible that other factors, such as maternal dietary polyphenol intake, may have led to transient changes in human milk flavonoid concentrations. Additionally, substantial variability in flavonoid concentrations were observed in both of these cohorts, which suggests that other factors likely play a role in modifying the qualitative and quantitative profiles of these compounds in human milk, ultimately impacting exposure to breastfed infants. Few studies have evaluated flavonoid metabolites in human milk across different stages of lactation, including Song et al. [52] and Khymenets et al. [28]. Using pooled samples from the previously described longitudinal, multinational cohort, Song et al. [52] carried out a preliminary evaluation of phase II flavonoid metabolites, including glucuronidated and methylated metabolites of epicatechin, epigallocatechin gallate, naringenin, kaempferol, hesperetin, and quercetin (**Fig. 5**). In general, concentrations of these metabolites were low in human milk (<10 nmol/L) and did not appear to be influenced by lactation stage. Khymenets et al. [28] detected and quantified phase II host epicatechin metabolites and microbial metabolites of flavan-3-ols (valerolactones) across different stages of lactation (colostrum, transitional, mature milk). Concentrations of the flavan-3-ol metabolites analyzed were not associated with lactation stage. However, evaluation of longitudinal changes was likely limited by the small sample sizes (colostrum, N = 3; transitional, N = 10; mature, N = 11). Concentrations of most metabolites were low (<40 nmol/L) and demonstrated little variability (13-39 nmol/L) and low detection in samples (detected in <30% of samples), with the exception of 5-(3ʹ-dihydroxyphenyl)-γ-valerolactone-4ʹ-sulfate (range: 20-8,305 nmol/L, detected in 92% of samples).

**Figure 4:**
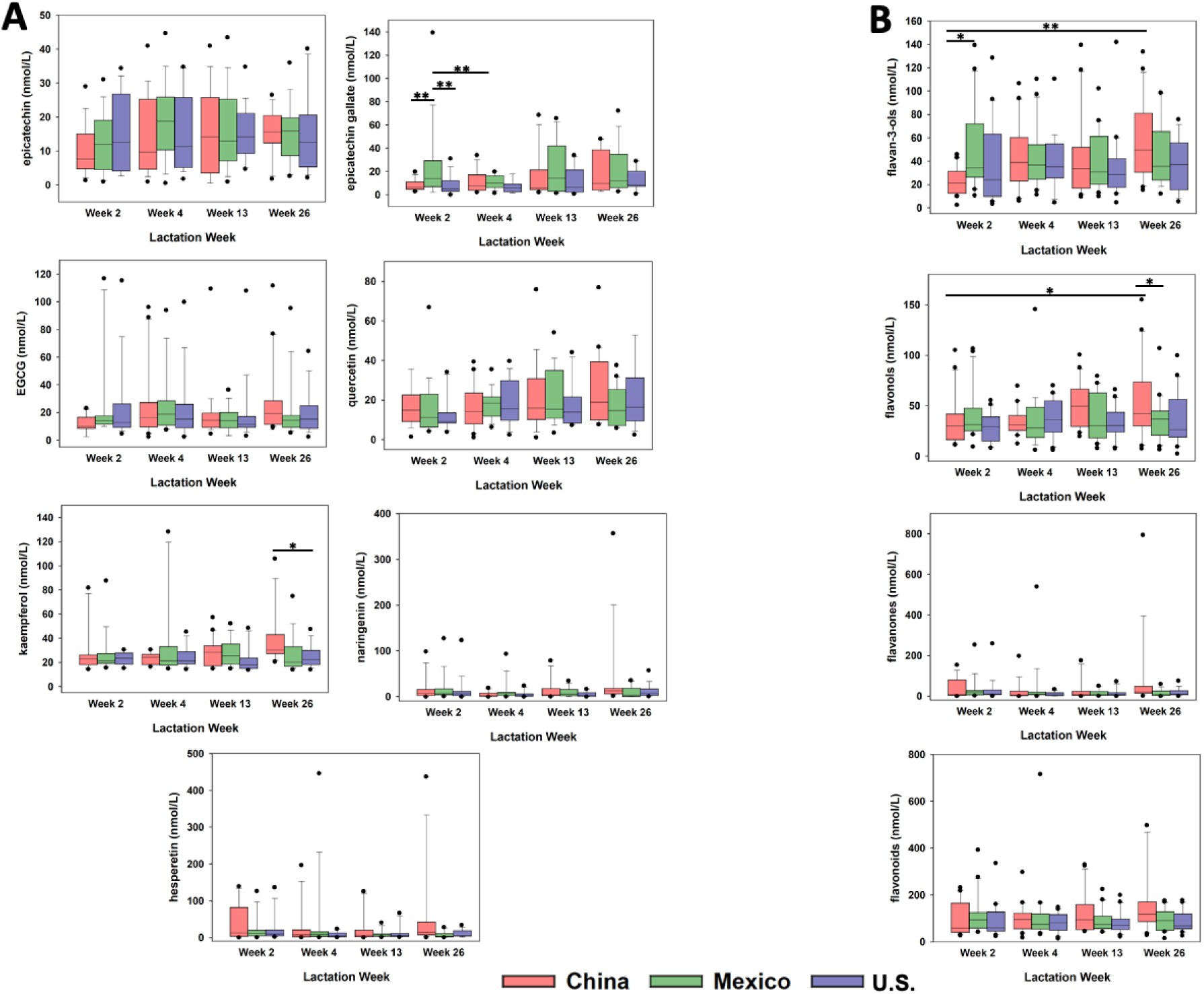
Boxplots showing differences in flavonoid concentrations (nmol/L) in human milk from a longitudinal, multinational cohort of lactating women from Mexico, China, and the United States (n = 20 per country) were adapted from Song et al. [52] with permission. Individual flavonoid concentrations (A) include flavan-3-ols (epicatechin, epicatechin gallate, and epigallocatechin gallate [EGCG]), flavonols (quercetin and kaempferol), and flavanones (naringenin and hesperetin). Flavonoid concentrations are summarized (B) as total flavan-3-ols, flavonols, flavanones, and flavonoids. Statistically significant differences are represented by * (*p* < 0.05) and ** (*p* < 0.01).

**Figure 5:**
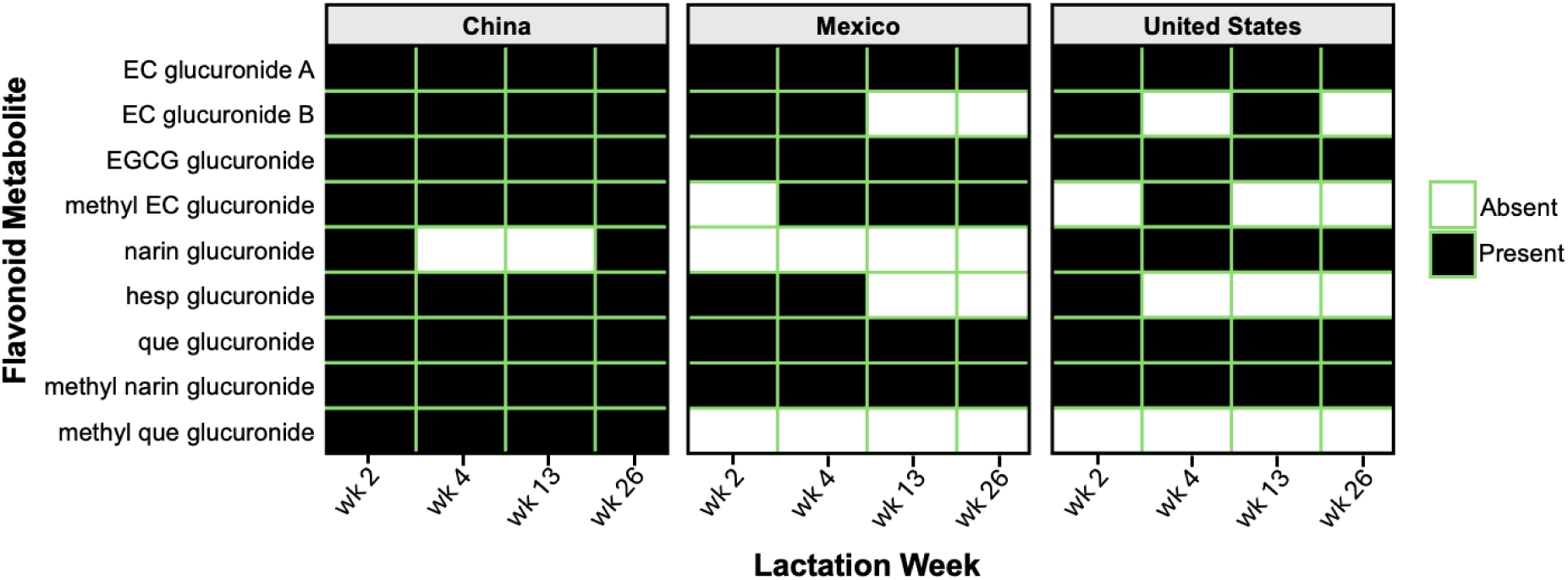
Presence and absence of phase II host flavonoid metabolites in human milk from a longitudinal cohort of lactating women from Mexico, China, and the U.S. (n = 20 per country) was adapted from [52] with permission. Presence and absence of each metabolite are represented by two pooled samples from each country by week combination with duplicate analyses. Boxes highlighted in green represent flavonoid metabolites that were detected in both replicates. EC, epicatechin; EGCG, epigallocatechin gallate; narin, naringenin; hesp, hesperetin; que, quercetin.

Sánchez-Hernández et al. reported a broader characterization of polyphenols in human milk, including various phenolic acids, flavonoids, and hydroxybenzaldehydes across three different lactation stages (colostrum, transitional, mature) [30]. In their analysis, only 2-vanillin, which is used a flavoring agent in food [105] and can also be generated as a gut microbial metabolite of phenolic acid precursors [106], was significantly associated with lactation stage, with greater concentrations in transitional milk (398.29 nmol/L) compared to colostrum (105.82 nmol/L). However, as with prior reports, this study was limited by small sample sizes (N = 7 per lactation stage), lack of data on maternal dietary intake, and substantial variability in concentration ranges. Overall, across the studies evaluating associations with lactation stage, there is a general lack of associations for most polyphenols and associations that have been observed are inconsistent. This may be related to the substantial degree of variability in polyphenol concentrations, which likely resulted from the non-standardized collection methodologies that do not account for the pharmacokinetics of these compounds relative to dietary intake. In addition, none of the studies that evaluated longitudinal dynamics of polyphenol profiles and concentrations in human milk controlled maternal dietary intake, which is a critical determinant of human milk polyphenol composition [13,14,28]. Collection strategies in these studies were also variable, ranging from a lack of standardization of extraction and analysis methodologies [28] to a lack of standardized collection timing and volume [27,30,52]. Interestingly, the studies with more standardization of collection timing and volume demonstrated that select polyphenols associate with lactation stage, although the observations were not consistent between studies. This may reflect differences in overall dietary polyphenol intake and timing of collection relative to prior dietary polyphenol intake. For example, conjugated flavonoid metabolites (e.g., sulfated or glucuronidated flavan-3-ols) have very short half-lives in circulation following dietary flavonoid intake. The half-lives in circulation for these metabolites is typically less than 3 hours [91]. Likewise, peak concentrations observed in human milk are typically reached within 3-6 hours [28]. Polyphenol metabolites derived from microbial metabolism, such as valerolactones or phenolic acids, have much longer half-lives, in some cases 24-48 hours [91]. Interestingly, Sánchez-Hernández et al. [30] utilized a 24-hr collection and, despite a small sample size, demonstrated that concentrations of the microbial metabolite 2-vanillin were associated with lactation stage. Future studies should include standardized collection timing (e.g., same time of day or 24-hr collection) and volume (e.g., full breast expression), while considering the pharmacokinetics of these compounds by: i) targeting parent polyphenols and conjugated metabolites within 3-6 hours of last dietary intake and ii) targeting microbial metabolites with 24-hr sample collection

#### 3.2.3. Maternal demographics and polyphenol profiles of human milk

Evaluations of polyphenol profiles in human milk have been carried out in a wide range of populations, including the U.S. [13,14,27,52], Spain [28–30], Germany [44], China [10,47,49,52], Poland [45,46,51], and Mexico [52]. However, there is only a single report of polyphenol profiles in a multinational study, which included cohorts of lactating women from the U.S., Mexico, and China [52]. In this analysis, Song et al. characterized profiles of flavonoids and phase II metabolites of flavonoids (**Fig. 4** and **Fig. 5**) and demonstrated cohort-specific (i.e., country-specific) differences in human milk flavonoid profiles. Specifically, differences in concentrations of epicatechin gallate, kaempferol, total flavan-3-ols, and total flavonols were observed at week 2 and week 26 postpartum. At week 2, the cohort from Mexico had the highest concentrations of epicatechin gallate and total flavan-3-ols. At week 26, the cohort from China had the highest concentrations of kaempferol and total flavonols. In a preliminary evaluation of flavonoid metabolites in this multinational cohort, the presence and absence of specific metabolites were consistent with differences in maternal demographics. Methyl epicatechin glucuronide was more consistently detected in the cohorts from Mexico and China compared to the U.S. Naringenin glucuronide was more common in the cohort from the U.S., while hesperetin glucuronide and methyl quercetin glucuronide were more consistently detected in the cohort from China. Expanding on these observations with deeper phenotypic characterization would provide insight into whether the differences associated with participant demographics is mediated by other dietary or non-dietary factors (i.e., genetics, health status, metabotype, microbiome). Pairing these data with a broader number of analytes and sample size would also facilitate exploration of factors responsible for the substantial degree of variability observed in the flavonoid concentration ranges observed in these cohorts. Dietary and supplement intake is a critical factor that was not evaluated in this multinational cohort. Substantial differences in dietary polyphenol intake exist between different countries. For example, average intakes in U.S. adults range between 1,200-2,000 mg/d [72], while intakes in Greece [107] and Mexico [108] can be as low as 600-800 mg/d. Dietary intake is the key driver of circulating profiles of polyphenols and phenolic metabolites [91,109] that are subsequently transferred from circulation to human milk. Therefore, it will be important for future studies evaluating differences in profiles of these compounds in human milk to pair these data with the participants’ dietary polyphenol intake. Gut microbial metabolism is also an important driver for the bioavailability of dietary polyphenols, with the majority of polyphenol metabolites reaching circulation being microbial-derived [91]. Although geographic region is associated with differences in gut microbiome composition [110], it has been suggested that geographic region is a proxy for multiple lifestyle factors, including diet, and genetics [111]. Since it is estimated that up to 8% of the variability in gut microbiome composition is explained by genetics [112], it is likely that complex relationships exist between geographic region, diet, genetics, and gut microbiome composition. Therefore, future studies evaluating the relationship between geographic region and polyphenol profiles of human milk should simultaneously evaluate these factors.

#### 3.2.4. Maternal metabotype and polyphenol profiles of human milk

The extensive variability in health outcome responses to dietary polyphenols has been hypothesized to be linked to variability in gut microbial structure (composition) and function [113,114]. While still needing to be mechanistically explored, this variability is throught to influence patterns of certain microbial metabolites in circulation that are produced from dietary polyphenol intake [115]. For example, pomegranate and walnuts are concentrated sources of ellagitannins, which are metabolized to urolithins by the gut microbiota [116]. *Gordonibacter* is associated with the production of specific urolithins [117], explaining, at least in part, the presence of urolithin metabotypes. Individuals have been categorized as metabotype 0 (urolithin non-producer), A (3,8-dihydroxy-urolithin producer), or B (produce combinations of 3,8- dihydroxy-urolithin, 3-hydroxy-urolithin, and 3,9-dihydroxy-urolithin) [118]. Cortés-Martín et al. demonstrated that urolithin concentrations in human milk were associated with maternal urolithin metabotypes with differential profiles in women categorized as urolithin metabotype A (e.g., 8-hydroxy-urolithin-3-glucuronide) vs. metabotype B (e.g., various urolithin glucuronides and sulfates) [29]. This demonstrates that maternal metabotype may be associated with differential profiles of urolithins in human milk, possibly due to differences in gut microbiome composition [119]. However, it is currently unknown whether metabotype is static or altered by changes in habitual dietary intake. Longitudinal studies are needed to evaluate relationships betweenthe dynamics of dietary changes, maternal urolithin metabotype, and human milk urolithin profiles.

Although dietary polyphenols are known to beneficially modulate gut microbiome composition in adults [120], the extent to which phenolic metabolites link to gut microbiome composition in breastfed infants is not yet known. Two pilot studies have demonstrated associations between polyphenol composition of human milk and gut microbiome composition in breastfed infants [13,29]. Cortés-Martín et al. showed that differential urolithin profiles in human milk, based on maternal urolithin metabotype, were associated with differences in *Gordonibacter* colonization during the first year of life [29]. Henning et al. demonstrated an association between the concentration of urolithin-3-glucuronide in human milk and relative abundance of *Blautia*, a butyrate-producer [13]. Factors driving infant gut microbiome composition are complex and include maternal gut and vaginal microbiome composition [121], delivery mode [121], preterm birth [122], maternal health status including diet [123], and infant dietary exposures (e.g., oligosaccharides and microbiota in human milk) [121,124,125]. To date, only one study has incorporated any of these factors into the evaluation of relationships between polyphenols in human milk and infant gut microbiome composition [29]. In a study conducted by Cortés-Martín et al. [29], it was shown that delivery mode did not impact the relationship between urolithin composition of human milk and *Gordonibacter* colonization. However, future studies are needed to determine whether delivery mode or any of these other factors might influence potential relationships between human milk polyphenols and infant gut microbiome composition. Integeating these factors into longitudinal studies evaluating both breastfed infant polyphenol intake and gut microbiome composition would provide valuable insight into understanding the extent to which polyphenol composition of human milk impacts infant gut microbiome development.

#### 3.2.5. Maternal diet and polyphenol profiles of human milk

Of the maternal factors evaluated, the potential influence of maternal dietary polyphenol intake on profiles of human milk polyphenols is the most well-studied but is limited to several short dietary interventions (≤ 4 weeks) [13,14,29,43,44,48] and single dose or dose escalation pharmacokinetic studies [28,29,42,45–47]. The types of analytes that have been quantified within this context includes flavonoids [44,48] and conjugated flavonoid metabolites [48], isoflavones [42,44,47], phenolic acids [48], urolithins [13,29,43,48], and valerolcatones [28]. In total, 49 analytes have been quantified (**Table 8** and **Table 9**) across these 11 studies, with 28 of these analytes quantified for the first time recently by our group [48]. In comparison, a combined total of 82 analytes have been quantified across ten studies in cohorts of free-living women [10,27,28,30,49–54], again with one recently published analysis representing a large proportion (34%) of the total [54]. Overall, increasing maternal dietary polyphenol intake has been shown to be associated with increased concentrations of polyphenols or related metabolites in human milk [13,14,28,29,42–44,46–48], with few studies that have failed to show a significant change in human milk polyphenol concentrations [44,45]. The discrepancy in results may be related to methodological differences such as sample collection and storage, extraction methodologies, or other factors influencing polyphenol metabolism that were not evaluated in these studies such as health status, gut microbiome composition, and genetics of host metabolism enzymes.

**Table 8.**
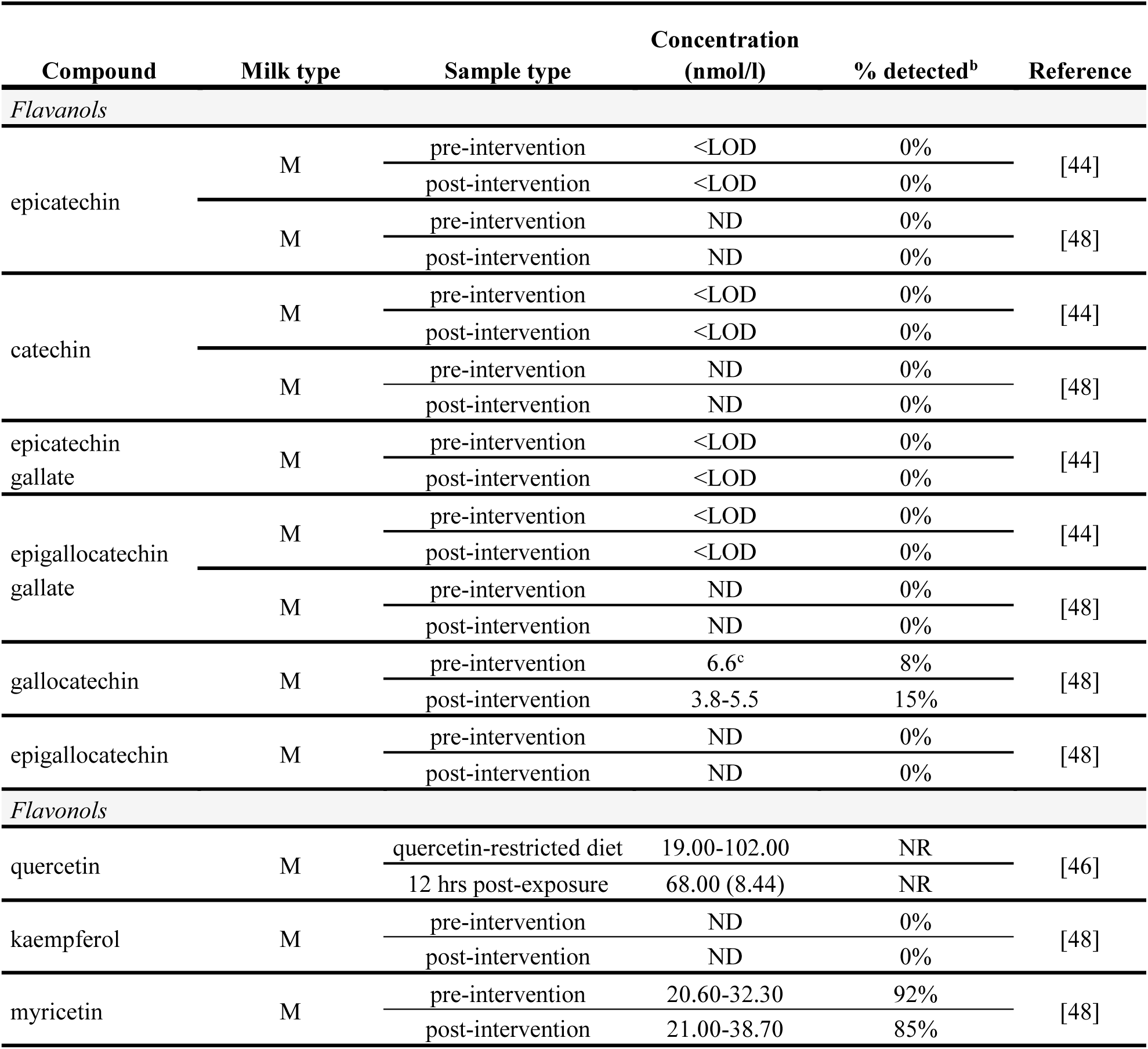
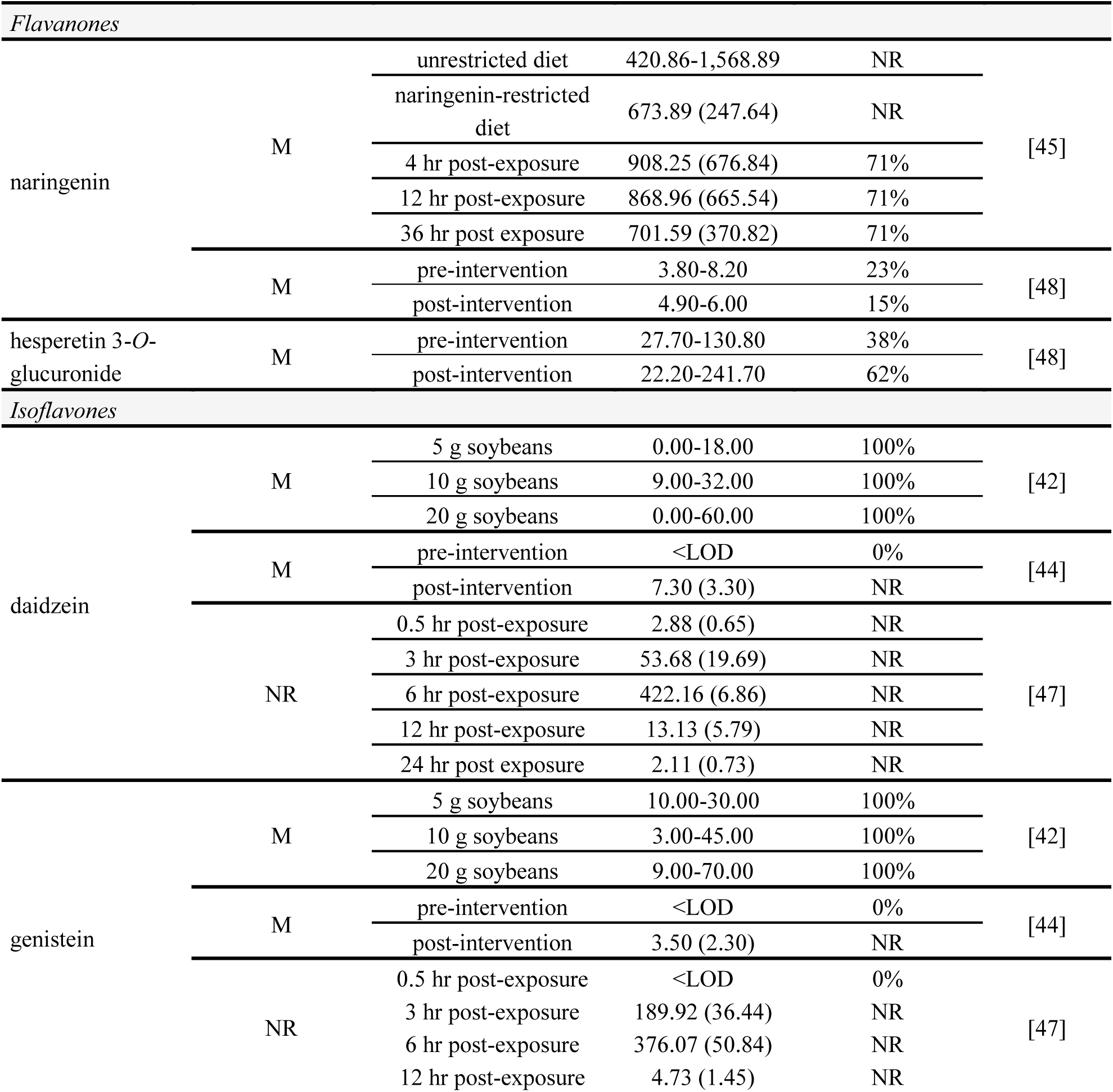
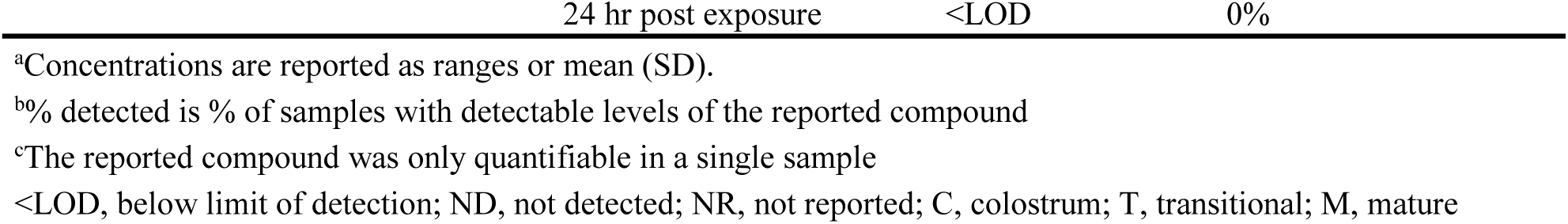
Flavonoids and flavonoid metabolites from food-derived sources detected and quantified in human milk in single dose and dietary intervention studies^a^.

**Table 9.**
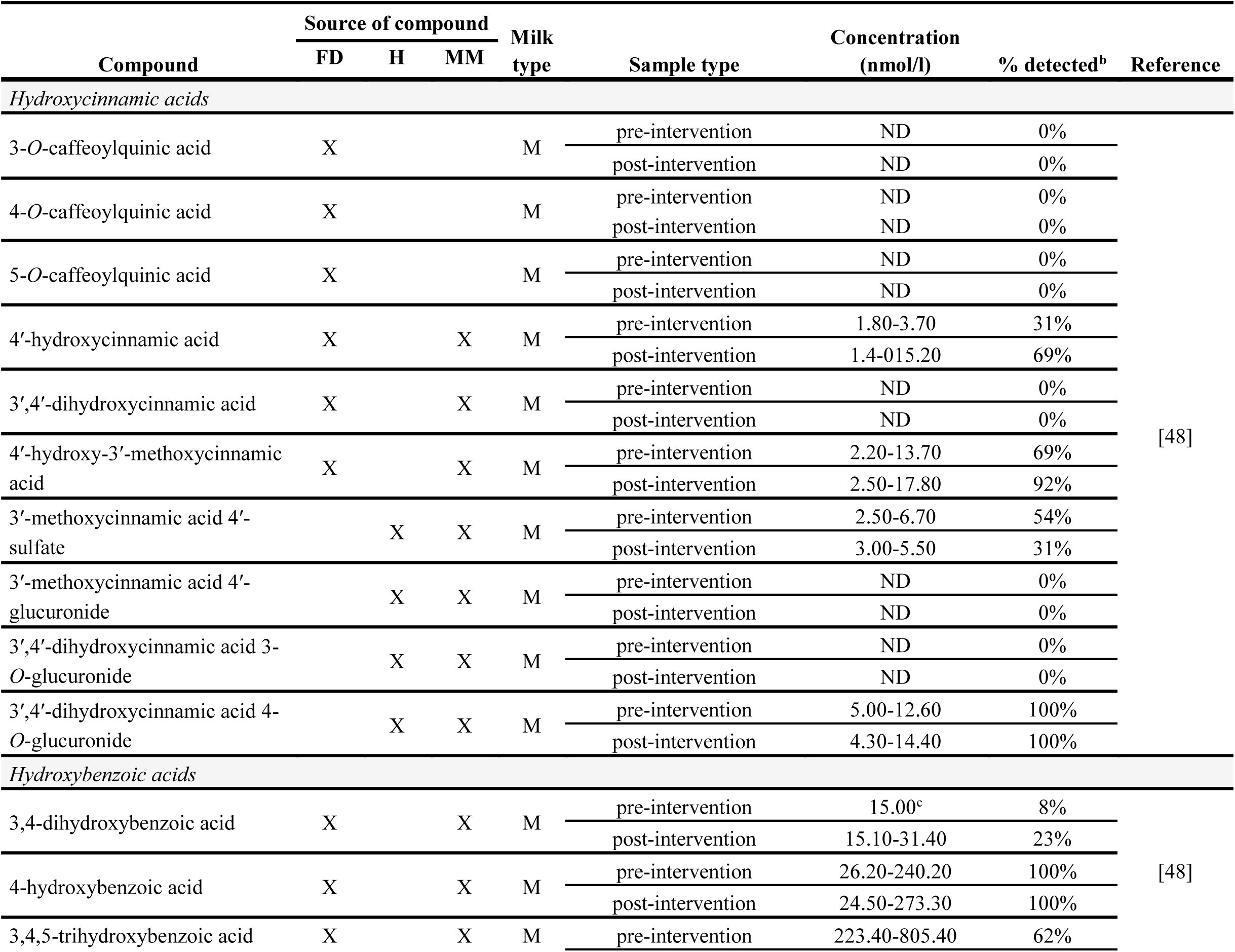
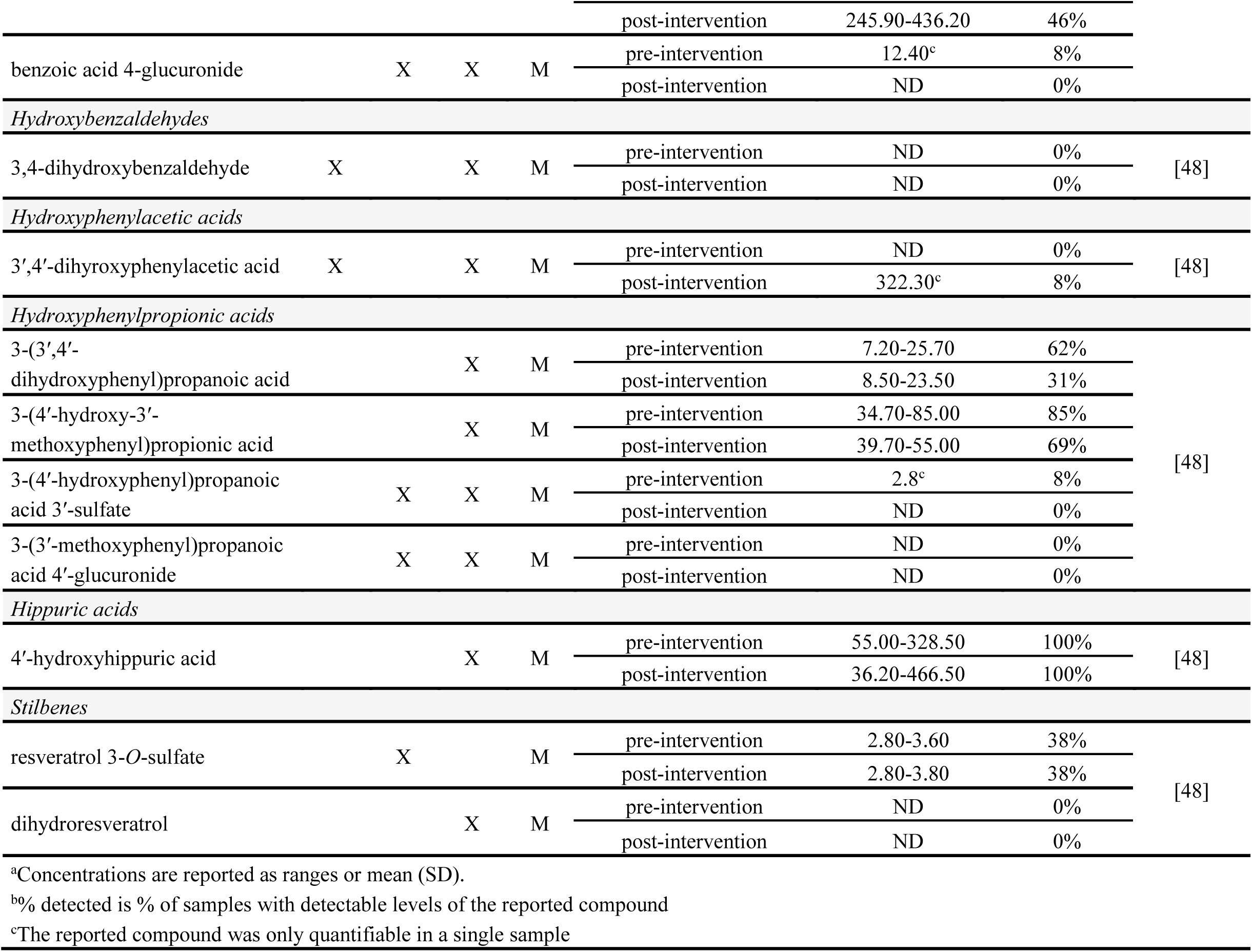
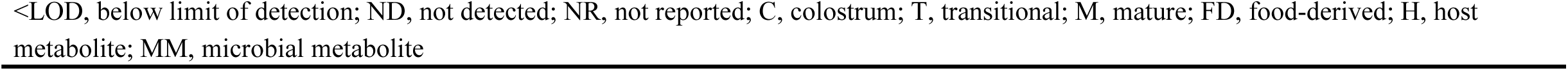
Phenolic acids and other phenolic compounds detected and quantified in human milk in dietary intervention studies^a^.

##### Maternal diet and flavonoids, flavonoid host metabolites, and microbial phenolic metabolites in human milk

Inconsistent relationships between maternal flavonoid intake and concentrations of flavonoids or flavonoid metabolites in human milk have been reported. In evaluations of flavonoid pharmacokinetics in human milk, Romaszko et al. [46] and Khymenets et al. [28] demonstrated increases of quercetin and phase II conjugates of flavan-3-ols, respectively. Romaszko et al. [46] evaluated the pharmacokinetics of quercetin in human milk after a single dose of onion soup (1 mg quercetin/kg body weight). The C_max_ for quercetin in human milk was 68 ± 8 nM at ∼12 hours post-exposure, which was a significant increase from baseline (40 nM). The quercetin concentration in human milk returned to baseline (42 nM) by 48 hours post-exposure. Khymenets et al. [28] examined pharmacokinetics of phase II conjugates of flavan-3-ols in human milk following consumption of dark chocolate (80 mg flavan-3-ols). Concentrations of these metabolites in human milk increased from below the limit of detection (LOD) at baseline to a C_max_ of 20-40 nM at 4-6 hours post-exposure and remained elevated above baseline levels even at 12 hours post-exposure (10-15 nM). This suggests that pharmacokinetics of flavan-3-ol metabolites may exhibit ∼2 hour delay in human milk relative to plasma (T_max_: 1-3 hours) [91,109].

Some studies have been unable to demonstrate the transfer of dietary flavonoids to human milk. For example, in a small pilot study of free-living lactating women (N = 24), Khymenets et al. [28] did not find significant associations between maternal flavan-3-ol intake determined from 3-day diet records and concentrations of 5-(3ʹ-dihydroxyphenyl)-γ-valerolactone-4ʹ-sulfate, a phase II conjugate (gut-host metabolite). This could be due to the small sample size, lack of consideration for the pharmacokinetics of this compound, or inaccuracies associated with self-reported dietary intakes and food composition databases. The unconjugated form of this metabolite, 5-(3’-dihydroxyphenyl)-γ-valerolactone, has a long half life (39.5 ± 26.4 hours) [91] and can persist at elevated concentrations in plasma for over 24 hours [109]. Thus, it is possible that targeting dietary intake during the 24-72 hour time period prior to milk collection may provide a more targeted evaluation of the relationship between dietary intake and valerolactones in human milk.

Likewise, Lu et al. [10] demonstrated that flavonoid intake in lactating women from Hong Kong (N = 89) was not associated with epicatechin concentrations in human milk. However, flavonoid intake did positively correlate with kaempferol, 3ʹ,4ʹ-dihydroxycinnamic acid, and 4ʹ- hydroxy-3ʹ-methoxycinnamic acid concentrations in human milk. This study suggests that microbial metabolites (e.g., phenolic acids) may correlate with maternal flavonoid intake, but the results should be interpreted with caution due to the lack of multiple test corrections (e.g., false discovery rate [FDR]) for the correlation analyses. In addition, more granularity in the dietary intake from this study would be needed to determine whether individual dietary flavonoids (e.g., kaempferol) directly correlate with concentrations of these compounds in human milk.

The study carried out by Lu et al., like that of Khymenets et al., did not take into account the long elimination kinetics of polyphenol metabolites when exploring the relationships between maternal diet and compounds such as epicatechin or phenolic acids in the milk. The timing of dietary intake records relative to milk collection should be determined based on the pharmacokinetics of these metabolites. For example, evaluating microbial metabolites such as phenolic acids could require multiple milk sampling time points during the 24-72 hours after recorded dietary intake.

Other reports [44,45] failing to chararacterize changes in flavonoid concentrations in human milk after increasing maternal flavonoid intake likely resulted from other study design limitations. For example, Romaszko et al. [45] evaluated the pharmacokinetics of the flavanone naringenin in human milk following intake of 250 ml of red grapefruit juice, which provided only 1 mg naringenin and 14 mg naringin. Participants consumed a naringenin-restricted diet for 5 days prior to consumption of the study product, but baseline concentrations of naringenin in human milk were reported to be 674 ± 248 nM, suggesting a lack of adherence to the dietary restrictions as significant dietary sources of naringenin were most likely still being consumed due to the short half-life of naringenin. Although this represents a decrease from concentrations prior to the naringenin-restricted diet (823 ± 285 nM), the reduction is quite modest (∼20%). The C_max_ occurred at ∼4 hours post-exposure with average concentrations of 908 ± 666 nM. Due to the substantial variability in naringenin concentrations, the increase observed was not statistically significant. It is possible that the narignenin concentrations reported by Romaszko et al. occurred due to the lack of washout. However, plasma concentrations of naringenin have been reported to reach <100 nM following a 40 mg dose of naringenin [126]. Thus, it is unlikely that human milk concentrations of naringenin would reach the concentrations reported by Romaszko et al. [45].

The results presented should be interpreted with caution as most of the concentrations reported in the study were below the assay limit of quantitation (LOQ, 1,469.18 nM). Jochum et al. carried out a short-term, 6-day dietary intervention of catechins from decaffeinated black tea (67 mg catechins/day) to evaluate the change in flavan-3-ol concentrations in human milk [44]. Over the course of the study, catechin concentrations remained below detectable levels. This may be due to the timing of the milk collection, which occurred ∼10 hours after intake. The extraction methods utilized allowed the total of catechins and phase II conjugates of catechins to be quantified. Peak concentrations of these compounds occur at 1-3 hours in plasma [91,109] and 4-6 hours in human milk [28]. Thus, collecting milk at 10 hours after intake could have resulted in the concentrations of these compounds falling below the LOD. Perhaps measuring microbial catechin metabolites, which have a longer half-life [91], would have yielded correlations with differences in dietary catechin intake.

Likewise, our recent evaluation of a 4-week polyphenol-rich healthy dietary intervention demonstrated that most flavan-3-ols in human milk fell below the LOD [48]. This may have resulted from the neutral pH of human milk [127], which is known to enhance oxidation of flavan-3-ol monomers [128]. We also evaluated concentrations of kaempferol, myricetin, naringenin, and hesperetin 3-*O*-glucuronide, none of which were influenced by the dietary intervention, possibly due to the short half life of these compounds.

##### Phenolic acids

Within the context of a dietary intervention, our group’s recent pilot study [48] is the first evaluation of the influence of maternal polyphenol intake on phenolic acid profiles and concentrations in human milk. This study was also the first to integrate a non-dietary factor, maternal α1,2-fucosyltransferase 2 (FUT2) phenotype, which is known to influence composition of HMOs [129,130]. FUT2 phenotype is also associated with differences in gut microbiome composition that could modify profiles of polyphenols in human milk due to the established link between gut microbiome composition and circulating polyphenol metabolites [91,113,114]. In our pilot study, we demonstrated that the influence of maternal diet on increasing 4ʹ-hydroxy-3ʹ-methoxycinnamic acid, 4ʹ-hydroxycinnamic acid, and 4-hydroxybenzoic acid concentrations was differentially associated with maternal FUT2 phenotype [48]. This suggests that consideration of non-dietary factors, such as genetics and gut microbiome composition, known to influence polyphenol absorption and metabolism is necessary to evaluate the relationship between maternal diet and polyphenol composition of human milk. Other consistently quantifiable phenolic acids in our pilot study that are derived from dietary intake included 3ʹ,4ʹ-dihydroxycinnamic acid 4-*O*-glucuronide, 3,4,5-trihydroxybenzoic acid, 3-(4ʹ-hydroxy-3ʹ-methoxyphenyl)propanoic acid, and 4-hydroxyhippuric acid. However, associations with dietary intake did not reach statistical significance, likely because of high variation in concentrations. Most of these analytes demonstrated substantial variability in concentration ranges, suggesting that future studies need to incorporate a more detailed analysis of maternal diet composition and non-dietary factors (e.g., health status, gut microbiome composition) to understand key drivers of phenolic acid profiles and concentrations in human milk.

##### Urolithins

Urolithins are microbial metabolites of ellagitannins and consistently increase in human milk in response to increased maternal dietary intake of ellagitannins derived from pomegranate or walnuts [13,29,43]. Cortés-Martín et al. [29] evaluated the total concentration of urolithins in human milk (N = 7) over 72 hours of daily walnut intake (30 g/day; up to 480 mg/d ellagitanins [131]) and characterized urolithin profiles in human milk (N = 27) after 72 hours of daily walnut intake (30 g/day). Total urolithin concentrations reached ∼28 nM at 72 hours and consisted primarily of 8-hydroxy-urolithin-3-glucuronide, 8-hydroxy-urolithin-3-sulfate, 9-hydroxy-urolithin-3-glucuronide, and urolithin-3-glucuronide. Henning et al. [43] carried out a proof-of-concept study in which lactating women (N = 2) consumed 8 oz. of pomegranate juice per day for two weeks (∼400 mg/d ellagitannins [132]). The concentrations of 8-hydroxy-urolithin-3-glucuronide increased from near or below the LOD to over 20 nM by day 14.

Henning et al. [13] expanded this to a small pilot study (N = 10) that also demonstrated an increase in 8-hydroxy-urolithin-3-glucuronide following two weeks of daily pomegranate juice intake (8 oz./day). Six hours after intake, concentrations were undetectable for all except one participant (0.6 nM), and increased to 3.2-28.7 nM by day 14 for all participants. These studies suggest that urolithin concentrations in human milk depend, in part, on maternal intake of ellagitannins. However, total recovery of urolithins in human milk is needed to directly evaluate the impact of maternal pomegranate juice (i.e., ellagitannin) intake on urolithin exposures for breastfed infants. Our group also evaluated 3,8-dihydroxy-urolithin, 3,9-dihydroxy-urolithin, and 9-hydroxy-urolithin-3-glucuronide concentrations in human milk before and after a polyphenol-rich dietary intervention during lactation [48]. However, the small number of samples in which urolithins were detected limited our ability to identify a relationship between maternal dietary intake and concentrations of these ellagitannin metabolites. Cortés-Martín et al. [29] demonstrated that pre-intervention concentrations of these analytes is generally low. Since our pilot study was not specifically designed to increase ellagitannin intake, it is likely that dietary ellagitannin intake was not sufficient to consistently increase urolithin concentrations to consistently detectable levels.

##### Isoflavones

A positive relationship has been consistently shown between maternal dietary isoflavone intake and isoflavone concentrations in human milk [10,14,42,44,47]. In a pilot study (N = 2), Franke et al. [42] demonstrated that the concentrations of the isoflavones daidzein and genistein in human milk increased in a dose-dependent manner. The time to reach maximum concentrations (t_max_) and return to baseline also depended on the amount of dietary isoflavone intake. The maximum concentrations reached ∼70 nM and ∼60 nM for genistein and daidzein, respectively. Franke et al. [14] also demonstrate that consumption of ∼55 mg of isoflavones/day as a soy protein beverage over 2-4 days (N = 18) resulted in ∼14-fold increase in total isoflavone concentrations in human milk (baseline: 5.1 ± 5.8 nM; after soy intake: 70.7 ± 50.8 nM).

Similarly, Jochum et al. [44] showed that consumption of 12 mg of isoflavones/day as a soy beverage over six days (N = 18) also increased concentrations of isoflavones in human milk. Daidzein and genistein concentrations were undetectable at baseline and reached 9.1 ± 4.8 nM and 4.3 ± 2.7 nM, respectively, on day 7. Zhou et al. [47] carried out a pharmacokinetic study (N = 6) of daidzein and genistein concentrations in human milk following a single dose of 500 ml of soy milk containing 20 mg isoflavones. Maximum concentrations (C_max_) of daidzein (422 ± 7 nM) and genistein (376 ± 51 nM) were reached at ∼6 hours after intake. Likewise, Lu et al. [10] demonstrated in a cohort of lactating women from Hong Kong (N = 89), that maternal dietary isoflavone intake positively corelated with daidzein concentrations in human milk. These studies demonstrate that concentrations of isoflavones in human milk depend, at least in part, on maternal dietary isoflavone intake.

#### 3.2.6. Methodological influences in assessment of polyophenol metabolite profiles in human milk

Besides isoflavones, evaluation of polyphenols and polyphenol metabolite profiles in human milk is relatively recent, with the first report published in 2013 [27]. As a result, comparisons between different studies are challenging due to a lack of standardized methodologies in sample collection, storage, extraction, and analysis. Methods of sample collection vary widely between studies (**Table 10**), including timing of collection relative to previous dietary intake, timing of collection during the feed, and total volumes collected (e.g. specified volume or full breast expression), and, in some cases, specific methods for how samples were collected or stored are not fully described. Because polyphenols and polyphenol metabolites are primarily derived from maternal dietary intake, the pharmacokinetics of these compounds needs to be taken into consideration to standardize collection timing relative to prior dietary exposure, in order to characterize occurrence and kinetics in milk. For example, if researchers wish to explore the availability of the parent flavonoids or conjugated metabolites (e.g., glucuronidated flavan-3-ols), an appropriate strategy would be to target collections within 3-6 hours of dietary intake. However, targeting microbial-derived metabolites such as phenolic acids or valerolactones with longer elimination half lives may require sample collections over 24-72 hours.

**Table 10.**
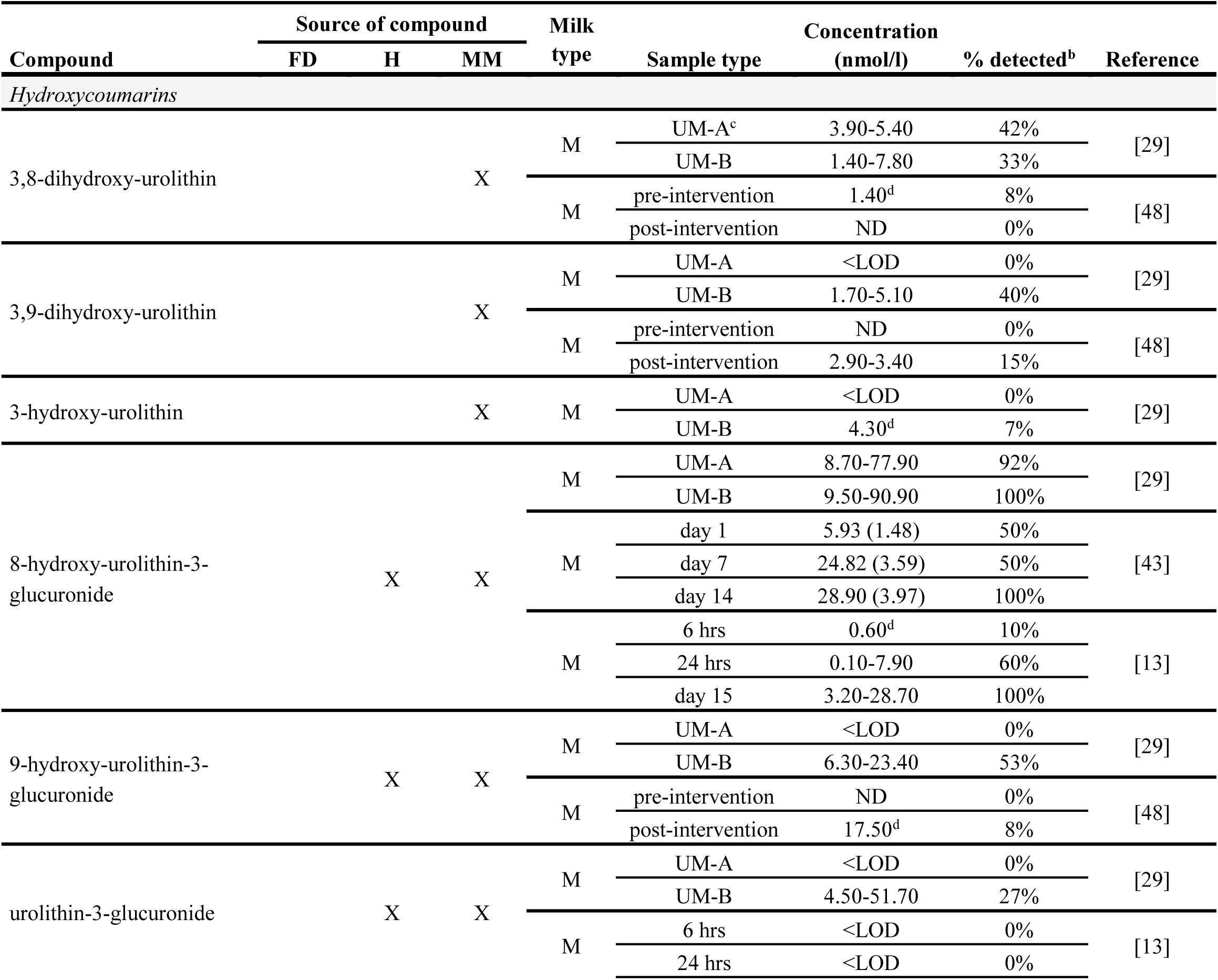
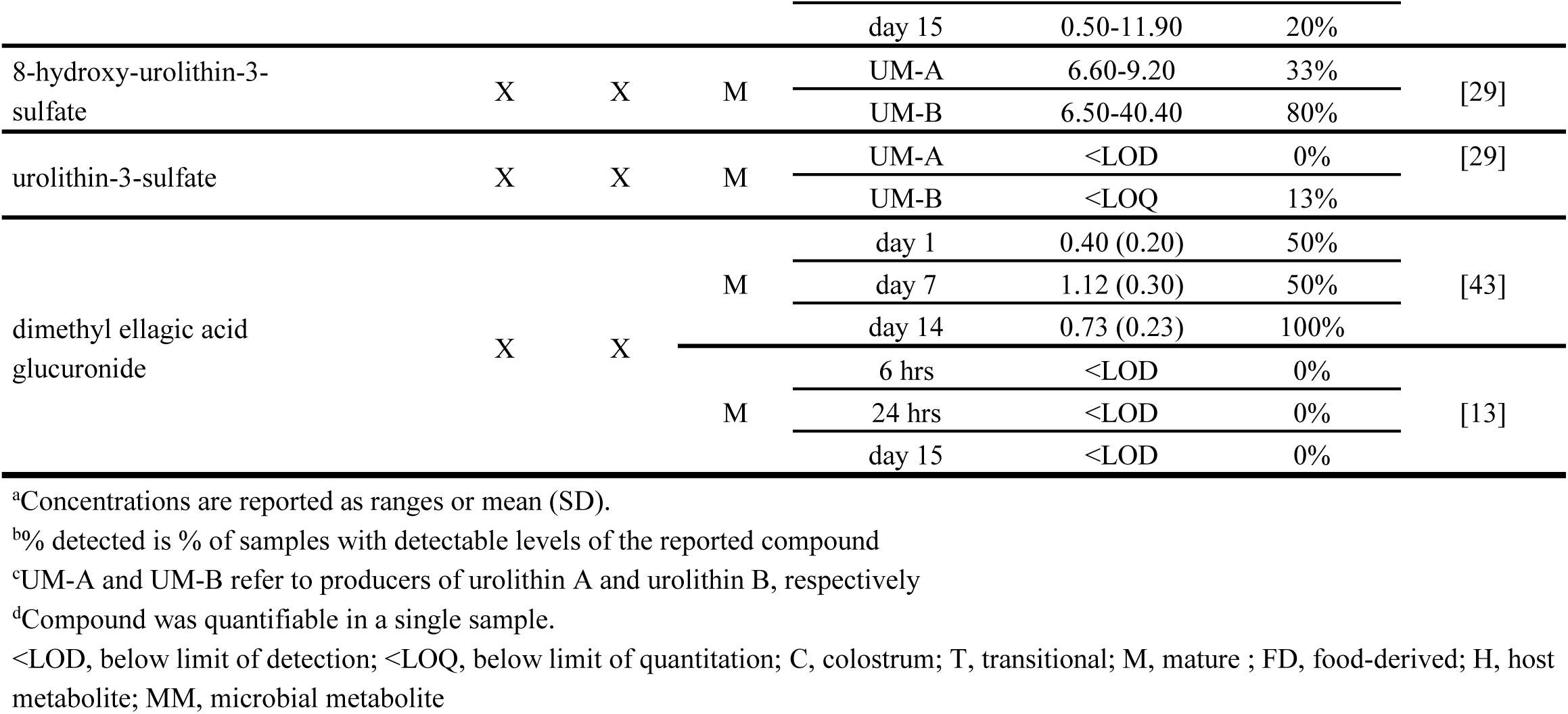
Ellagitannin metabolites detected and quantified in human milk in dietary intervention studies^a^.

Storage conditions between studies also varies, specifically the length of time that samples are stored at either 4 °C or −20 °C prior to cryogenic storage at −80 °C. Although evaluations of stability have been carried out for isoflavones in human milk [14,42], this has not been determined for other polyphenols or polyphenol metabolites. Even in the case of the isoflavones, published data describing stability under varying storage conditions have not been reported.

Stability of certain polyphenol subclasses, such as flavan-3-ols, is pH-dependent, with increased degradation occurring at neutral or slightly alkaline pH [133]. Therefore, ideal sample storage is to immediately freeze at −80 °C. However, in cases where this is not feasible, sample collection and storage methodologies need to be standardized to normalize storage conditions such as length of time when stored at home in a 4 °C refrigerator or −20 °C freezer.

Substantial differences in extraction and analytical methodologies also exist which affect the comparison of findings across studies. A major limiting factor is in the comparison of data derived from workflows which use acid or enzyme hydrolysis to those which do not. Hydrolysis is a method for removing the glucuronide or sulfate conjugates from the polyphenol backbone (aglycone), to reduce complexity of the numbers of metabolites measured. Although this practice is a simple way to establish total bioavailability, it limits the ability to characterize the phase II metabolites of polyphenols and their metabolites or compare findings with studies which do not use hydrolysis techniques. The majority of the published reports have analyzed the hydrolyzed polyphenol aglycone forms [10,14,27,42,44–47,49–53], with only eight evaluations of non-hydrolyzed polyphenols and metabolites [13,28–30,43,48,52,54]. Because analysis of the hydrolyzed aglycones would represent the total of the parent (precursor) compound (e.g., epicatechin) in addition to phase II conjugates (e.g., glucuronidated, and sulfated forms of epicatechin), this type of analysis may yield concentration ranges that differ from actual concentrations as hydrolysis efficiency is highly variable. As a result, it is challenging to compare concentration ranges of the studies that analyzed the hydrolyzed aglycone forms with those that evaluated the non-hydrolyzed compounds. About 60 individual polyphenols and related metabolites have been identified and quantified in human milk. However, estimating total concentrations of polyphenols or classes and subclasses of polyphenols remains a challenge due to the limited number of analytes included in individual assays, particularly those using hydorlysis methodology. Currently, the largest number of analytes to be evaluated simultaneously is 86, which was carried out by Berger et al. [54]. Now that a more substantial level of preliminary data exists on human milk phenolic profiles, a more complete and accurate evaluation of polyphenol metabolite exposures and intake by breastfed infants would require application of a broader targeted metabolomics assay and standardized collection and extraction methodologies.

### 3.3. Potential developmental impacts of polyphenol exposures in breastfed infants

#### 3.3.1. Absorption of polyphenols from human milk by breastfed infants

There is emerging clinical evidence demonstrating the transfer of polyphenols or related metabolites directly from human milk to breastfed infant circulation [13,14,49], including ellagitannin metabolites and isoflavones (**Table 11**). However, establishing a link between infant exposure and intake of polyphenol metabolites and clinical outcomes is limited by the availability of dietary intake data in breastfed infants and in lactating mothers. Currently, only concentrations in human milk and urine samples from breastfed infants have been quantified, but polyphenol intake by breastfed infants has not been estimated beyond milk concentration. Because bioactivity of dietary polyphenols has been linked to the concentrations of polyphenol metabolites in circulation in a dose-dependent manner in adults [31], it is possible that clinically relevant developmental impacts of polyphenol exposures from human milk could be identified. Besides direct effects of circulating polyphenol metabolites, some polyphenols or polyphenol metabolites (e.g., flavonoids) in human milk may have poor bioavailability [91] leading to developmental impacts associated with modulation of the gut microbiota of breastfed infants.

**Table 11.**
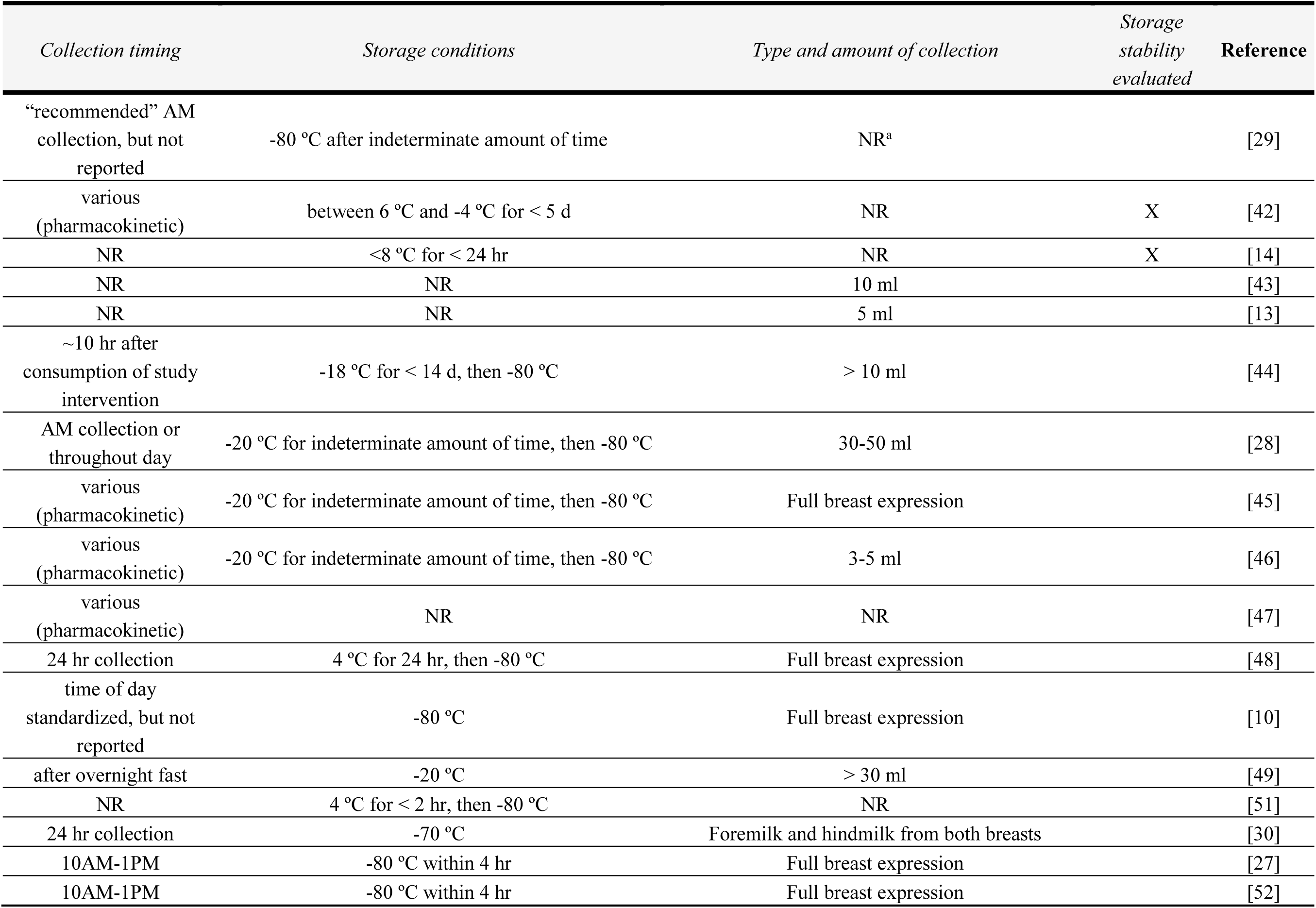
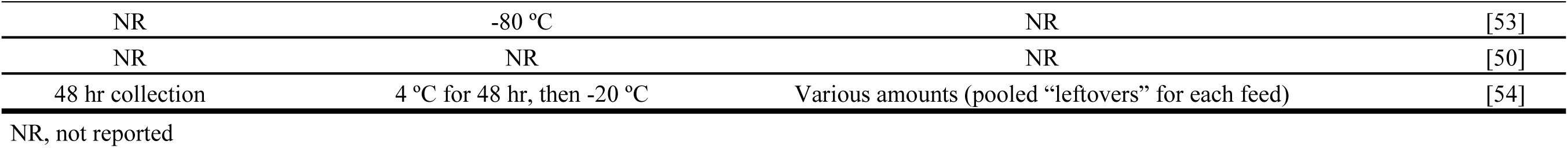
Human milk collection and storage conditions across different studies.

| *Collection timing* | *Storage conditions* | *Type and amount of collection* | *Storage stability evaluated* | **Reference** |
| --- | --- | --- | --- | --- |
| “recommended” AM collection, but not reported | -80 °C after indeterminate amount of time | NR<sup>a</sup> |  | [29] |
| various (pharmacokinetic) | between 6 °C and -4 °C for < 5 d | NR | X | [42] |
| NR | <8 °C for < 24 hr | NR | X | [14] |
| NR | NR | 10 ml |  | [43] |
| NR | NR | 5 ml |  | [13] |
| ~10 hr after consumption of study intervention | -18 °C for < 14 d, then -80 °C | > 10 ml |  | [44] |
| AM collection or throughout day | -20 °C for indeterminate amount of time, then -80 °C | 30-50 ml |  | [28] |
| various (pharmacokinetic) | -20 °C for indeterminate amount of time, then -80 °C | Full breast expression |  | [45] |
| various (pharmacokinetic) | -20 °C for indeterminate amount of time, then -80 °C | 3-5 ml |  | [46] |
| various (pharmacokinetic) | NR | NR |  | [47] |
| 24 hr collection | 4 °C for 24 hr, then -80 °C | Full breast expression |  | [48] |
| time of day standardized, but not reported | -80 °C | Full breast expression |  | [10] |
| after overnight fast | -20 °C | > 30 ml |  | [49] |
| NR | 4 °C for < 2 hr, then -80 °C | NR |  | [51] |
| 24 hr collection | -70 °C | Foremilk and hindmilk from both breasts |  | [30] |
| 10AM-1PM | -80 °C within 4 hr | Full breast expression |  | [27] |
| 10AM-1PM | -80 °C within 4 hr | Full breast expression |  | [52] |

|  |  |  |  |
| --- | --- | --- | --- |
| NR | -80 °C | NR | [53] |
| NR | NR | NR | [50] |
| 48 hr collection | 4 °C for 48 hr, then -20 °C | Various amounts (pooled “leftovers” for each feed) | [54] |
| NR, not reported |  |  |  |

**Table 12.** Phenolic compounds detected and quantified in urine of breastfed infants^a^.

| Study type and compounds quantified | Source of compound |  |  | Time point | Concentration (nmol/l) | % detected<sup>b</sup> | Reference |
| --- | --- | --- | --- | --- | --- | --- | --- |
|  | FD | H | MM |  |  |  |  |
| Dietary Intervention |  |  |  |  |  |  |  |
| 8-hydroxy-urolithin-3-glucuronide |  | X | X | pre (day 0) | <LOD | 0% | [13] |
|  |  |  |  | post (day 15) | 8.90-40.30 | 100% |  |
| 3-hydroxy-urolithin |  | X | X | pre (day 0) | <LOD | 0% |  |
|  |  |  |  | post (day 15) | 0-17.50 | 20% |  |
| dimethyl ellagic acid glucuronide |  | X | X | pre (day 0) | <LOD | 0% |  |
|  |  |  |  | post (day 15) | <LOD | 0% |  |
| total isoflavones<sup>c</sup> | X |  |  | pre | 29.80 (11.60) | NR | [14] |
|  |  |  |  | post | 186.10 (25.10) | NR |  |
| Free Living |  |  |  |  |  |  |  |
| genistein | X |  |  |  | 96.00 (16.00) | 100% | [49] |
| daidzein | X |  |  |  | 44.00 (95.00) | 72% |  |
| equol |  |  | X |  | 47.00 (104.00) | 66% |  |
| glycitein | X |  |  |  | 232.00 (354.00) | 32% |  |
| enterolactone |  |  | X |  | <LOD | 0% |  |

<sup>a</sup>Concentrations are reported as ranges or mean (SD).<sup>b</sup>% detected is % of samples with detectable levels of the reported compound<sup>c</sup>Concentrations reported from this study are in nmol/mg creatinine
<LOD, below limit of detection; NR, not reported; FD, food-derived; H, host metabolite; MM, microbial metabolite.

#### 3.3.2. Associations between polyphenols in human milk and the gut-brain axis

Associations between polyphenol metabolites in human milk, specifically urolithins, have been positively associated with the abundances of *Gordonibacter* (urolithin-producing bacterium) [29] and *Blautia* (butyrate-producer) [13] in the gut of breastfed infants. Infant gut microbiomes characterized by increased abundance of probiotic taxa are positively associated with the development of language and fine motor skills [134,135]. Preclinical studies suggest that neuroactive microbiota-derived tryptophan metabolites, including indole-3-acetic acid (IAA), indole-3-lactic acid (ILA), and indole-3-propionic acid (IPA), may influence cognitive development by modulating inflammatory responses and neuronal development [136–138]. Since cognitive development in breastfed infants has been linked to tryptophan metabolites such as ILA within the context of other human milk bioactives (e.g., HMOs), it is possible that similar modulation of the gut-brain axis may occur in response to intake of human milk polyphenols. In fact, clinical evidence in adult populations suggest that the relationship between dietary polyphenol intake and cognitive function [33] is associated with similar modulation of the gut-brain axis through positive impacts to both probiotic taxa [120,139] and production of IAA, ILA, and IPA [140–142]. Consistent with this, several groups have reported positive impacts of polyphenol-rich fruits and vegetables on neurodevelopment and cognition in infants and children, either from maternal intake during pregnancy or child intake [35,38,40]. In addition, a preclinical model reflective of dietary polyphenol intake in young children (1.5-3 years old) has demonstrated accumulation of flavonoid metabolites in the cerebellum and frontal cortex [143] at concentrations known to influence neuroplasticity [144]. Therefore, it is also possible that impacts of human milk polyphenols on cognitive development in breastfed infants could occur directly from polyphenol metabolites reaching circulation. However, future studies are needed to evaluate the relationship between human milk polyphenols and the gut-brain axis of breastfed infants.

## 4. Research priorities for future studies

There is substantial variability in qualitative profiles and concentration ranges reported for human milk polyphenols and related metabolites in existing literature. Currently, clear links between maternal dietary and non-dietary factors, profiles of polyphenols in human milk and breastfed infant intake, and clinically relevant developmental outcomes have yet to be established. To establish the presence or absences of these relationships, it is critical to improve our understanding of how human milk polyphenol concentrations and profiles are regulated and what factors contribute to infant exposure through breastfeeding. A crucial first step for future studies is to: 1) evaluate polyphenol intakes at the population level for lactating women, 2) standardize collection and storage to prevent polyphenol degradation, 3) standardize analytical methodologies and population sampling to capture a broader range of dietary intake and infant exposures, including quantification of parent polyphenols and polyphenol metabolites, and 4) evaluate polyphenol pharmacokinetics, including correlations to gastric emptying and intestinal transit time, as a key consideration for timing and duration of sample collection. Although inconsistencies in study design, sampling and analytical methods exist, diet, metabotype, and lactation stage have been identified as potential factors associated with differential human milk polyphenol profiles. These inconsistencies may reflect the absence of studies that have systematically integrated:

1. Broader assessment of polyphenols and polyphenol metabolites in both human milk and maternal diets
2. Maternal factors that may influence polyphenol metabolism, such as health status, gut microbiome composition, and genetics of host metabolism enzymes (e.g., catechol-*O*-methyltransferase), form of intake (e.g., beverage, food, supplementation), and polyphenol dose
3. Maternal non-dietary factors known to influence composition of other bioactive components in human milk, such as lactation stage, diet, and genetics

This type of systematic integration would allow clearer links to be established between maternal factors and human milk polyphenol profiles, but will require cohorts and intervention studies to have deeper phenotypic characterization, larger sample sizes and greater ethnic and genetic diversity. Extending this to link human milk polyphenol profiles to developmental outcomes (e.g., gut-brain axis) in breastfed infants will require: 1) quantification of infant dietary intake of individual polyphenols and their metabolites in human milk, 2) longitudinal evaluation of developmental outcomes, and 3) phenotypic characterization of factors that may influence polyphenol metabolism or outcomes of interest.

## 5. Summary

Polyphenols and polyphenol metabolites in human milk may play a role in modulation of the gut-brain axis of breastfed infants. However, there is a paucity of current evidence directly linking human milk polyphenol composition and/or intakes to developmental outcomes in breastfed infants. A deeper, systematic phenotypic characterization integrating 1) dietary and non-dietary factors, 2) broader polyphenol profiling of maternal diets and human milk, 3) quantification of polyphenol intakes in breastfed infants, and 4) developmental outcomes in breastfed infants is needed to establish that the transfer of polyphenols from maternal diets to breastfed infants positively impacts developmental outcomes.

## Data availability statement

The authors confirm that the data supporting the findings of this study are available upon request.

## Authors’ contributions

C.F.: Conceptualization, methodology, formal analysis, investigation, data curation, writing – original draft, writing – review & editing; N.K.: Formal analysis, data curation; A.A.: writing – review & editing; M.F.: Conceptualization, methodology, writing – review & editing, supervision; C.K.: Writing – review & editing, supervision.

